# Potent clinical predictive and systemic adjuvant therapeutic value of plasma fractalkine in PD-L1/PD-1 blockade immunotherapy for lung cancer

**DOI:** 10.1101/2022.06.16.22276511

**Authors:** Ana Bocanegra, Gonzalo Fernández, Daniel Ajona, Hugo Arasanz, Ester Blanco, Miren Zuazo, Luisa Chocarro, Sergio Piñeiro-Hermida, Pilar Morente, Leticia Fernández, Maider Garnica, Ana Remirez, Maite Martinez-Aguillo, Idoia Morilla, Beatriz Tavira, Pablo Ramos, Miriam Echaide, Juan José Lasarte, Luis Montuenga, Ruth Vera, Ruben Pio, David Escors, Grazyna Kochan

## Abstract

Recent studies highlight the importance of baseline functional immunity for efficacious immune checkpoint blockade therapies. High-dimensional systemic immune profiling was performed in a discovery cohort of 112 non-small cell lung cancer patients undergoing PD-L1/PD-1 blockade immunotherapy. Responders showed high baseline phenotypic diversity of myeloid cell types in peripheral blood, in which elevated activated monocytic cells and decreased granulocytic phenotypes were potent predictive biomarkers. High-throughput profiling of soluble factors in plasma identified fractalkine (FKN), a chemokine involved in immune chemotaxis and adhesion, as a biomarker of myeloid cell diversity in human patients and in murine models, which was found significantly increased in objective responders. Secreted FKN inhibited adenocarcinoma and squamous cell carcinoma growth in vivo through a prominent contribution of systemic effector NK cells, enhanced tumor infiltration with immunostimulatory immune cells and inhibition of MDSCs within tumors. A synergy between FKN and PD-L1/PD-1 blockade immunotherapy was found in murine lung cancer models refractory to anti-PD-L1/anti-PD-1 treatment. Transcriptional data from 515 human lung adenocarcinoma samples independently confirmed the results from the discovery cohort. Importantly, recombinant FKN and tumor expressed-FKN were efficacious in delaying tumor growth in vivo with significant abscopal effects, indicating a potential therapeutic use of FKN in combination with immunotherapies.

**One Sentence Summary:** Serum fractalkine as a biomarker of response to immune checkpoint blockade.

## INTRODUCTION

Non-small cell lung cancer (NSCLC) remains a leading cause of cancer death. PD-L1/PD-1 blockade immunotherapies have yielded remarkable clinical benefit and durable responses. Nevertheless, these treatments fail in most patients. The mechanisms of resistance to immune checkpoint blockade (ICB) therapies have traditionally been studied within the tumor microenvironment. However, PD-L1/PD-1 blocking antibodies are administered systemically, acting over a variety of systemic immune populations that may contribute to clinical outcomes (*1-3*). Accumulating evidence demonstrate the importance of functional systemic immunity in cancer patients before starting immunotherapies as a pre-requisite for ICB success (*1, 4-10*). Indeed, we and others have recently demonstrated that a functional baseline systemic T cell immunity is a major requirement for PD-L1/PD-1 blockade efficacy in cancer patients (*4, 6-8*). Patients with dysfunctional T cells failed to respond to PD-L1/PD-1 monotherapies and had increased risk of developing hyperprogressive disease (*11*). Broad immune cell profiling from routine clinical blood analyses has been classically studied as prognostic variables (*12*), which associate to better therapeutic outcomes independently of the treatments. Amongst these, neutrophil-to-lymphocyte ratio (NLR), and lymphocyte, monocyte and platelet counts are the most widely used. However, the applicability of these biomarkers largely depends on the specific analytical techniques applied for routine blood analyses in clinical practice.

We previously demonstrated that baseline T cell functionality in NSCLC patients was required for objective responses to PD-L1/PD-1 blockade monotherapies (*4*). T cell activities are ultimately regulated by cells of myeloid origin through antigen presentation. In turn, the differentiation of myeloid cell subsets and their activation status in cancer are systemically regulated by cytokines and plasma factors (*13*). These factors can potentially perturb myelopoiesis and alter the diversity of immune myeloid cell repertoires, and hence T cell activities. Thus, tumors that overexpress GM-CSF, G-CSF, M-CSF and IL-6 ultimately lead to systemic expansion of immunosuppressive MDSCs that counteract anti-tumor T cells and NK cells (*14-16*). In contrast, patients with increased HLA-DR^+^ monocytes and decreased neutrophils in peripheral blood show good response rates to ICB therapies (*17*). Specific cytokine profiles associated with different cancer types may alter systemic immunity at differing degrees, affecting the prognosis of ICB therapies. While some cytokines are clearly strongly immunosuppressive, others may be associated with good prognosis and could be used as biomarkers of response. Indeed, some of these cytokines, such as IL-2, IFNα or IL-12, may be administered in combination with ICB to reinvigorate immunoreactivity (*18*).

Here, we have performed an extensive profiling of systemic myeloid cell populations in NSCLC patients undergoing PD-L1/PD-1 blockade therapies, before and during treatment. Responder patients showed an elevated diversity of myeloid cell types, enriched in activated monocytic cells and decreased granulocytic populations. Thus, a high-throughput characterization of plasma factor signatures was carried out to associate specific profiles with systemic immune cell diversity and objective responses. Plasma fractalkine (FKN, CX3CL1) concentration was the best candidate for a biomarker of response, which correlated with systemic myeloid diversity and response to PD-L1/PD-1 blockade.

FKN is a chemokine that belongs to the CX3C family. This protein is mainly expressed by endothelial cells and signals through its canonical receptor CX3CR1, and α_v_β_3_ and α_4_β_1_ integrins (*19*). It is produced as a membrane-bound protein that mediates cellular adhesion. A soluble form released by metalloproteases such as ADAM-10 and ADAM-17 (*20, 21*) regulates immune chemotaxis. FKN has been reported to elicit anti-tumor responses by recruiting T cells, dendritic cells (DCs) and natural killer (NK) cells to the tumor microenvironment (*22-30*). In contrast, FKN was shown to promote metastasis of CX3CR1^+^ circulating tumor cells towards tissues displaying a high CX3CL1 expression, such as bones, lungs and nervous tissues in several types of carcinomas, such as breast cancer (*31*). This pro-oncogenic role of FKN is attributed to its ability to promote tumor migration and invasion (*32-38*). Thus, FKN plays context-dependent roles in anti-tumor immunity.

Here, we explored *in vitro* and *in vivo* the underlying mechanisms explaining the beneficial contribution of FKN to ICB immunotherapy in NSCLC, and its use as a therapeutic agent.

This should include introductory information that lays out the clinical problem addressed by the research and that explains other background necessary for understanding the study.

## RESULTS

### Baseline diversity of myeloid cell subsets in peripheral blood is a signature of objective clinical responses

First, we evaluated classical variables based on standard clinical quantification of immune cell populations in peripheral blood of patients undergoing PD-L1/PD-1 blockade immunotherapy. These analyses were performed on a well-characterized cohort of 112 NSCLC patients **(Table S1)** and described in previous studies elsewhere (*2, 4, 11*). The majority of patients were smokers, 75% male and the mutational status of the tumors was not unknown in 96.4%. Hence, data were not stratified by sex or genetic alterations. Absolute counts of lymphocytes, monocytes, neutrophils and platelets before starting treatments were quantified. No significant differences were found between responders and non-responders after having analyzed routine clinical parameters, such as NLR, monocyte/lymphocyte ratio and platelet/lymphocyte ratio, or absolute monocyte and neutrophil counts **(Fig. S1)**, thus indicating their limited ability to predict response.

We then performed an extensive characterization of circulating immune cell populations by high dimensional flow cytometry (HDFC). This technique allows the simultaneous quantification of multiple cell lineage and activation markers in biological samples (*6, 17, 39*), leading to detailed identification of immune cell types and differentiation stages. A panel of 43 lineage, differentiation and activation markers was used to label freshly isolated peripheral blood mononuclear cells (PBMCs) from patients prior to the initiation of immunotherapy (baseline) and before the first cycle of immunotherapy. As controls, samples from age-matched healthy donors were used. Patient groups were classified into long-term responders, stable disease and progressors, with some of the progressors in our cohort classified as hyperprogressors in previous studies (*4, 11*).

High-dimensional hierarchical clustering was performed over the phenotypes of baseline CD11b^+^ myeloid immune populations. Each response group was characterized by distinct cluster profiles **(Fig. S2)**. Responders showed high phenotypic diversity with a dominance of the monocytic lineage, in contrast to non-responders who exhibited increased neutrophils and granulocytic myeloid-derived suppressor cells (G-MDSCs). A diversity index (DI) was calculated for each patient as the number of terminal phenotype clusters in HDFC profiles generated with 200,000 CD11b^+^ cells as an indicator of myelopoiesis proficiency, and correlated to responses (**Fig. 1A**). Objective responders exhibited increased (p<0.0001) baseline phenotypic myeloid diversity compared to non-responders before starting immunotherapy (DI=18.63 ± 2.36 vs 12.21 ± 3.31; Mean ± SD), although still inferior to that of healthy donors (DI=29.86 ± 3.71; Mean ± SD). Elevated diversity indexes were significantly associated with objective clinical responses as evaluated by ROC analysis. The area under the curve (AUC) was 0.9216 (95% confidence interval = 0.8601 to 0.9832; p<0.0001) (**Fig. 1B**). A DI cut-off value of 13.5 identified responders with 100% sensitivity and 63% specificity. This value stratified patients with significantly (p<0.0001) increased progression-free survival (PFS) (**Fig. 1C**) and overall survival (OS) (p=0.007) (**Fig. 1D**).

**Fig. 1.**
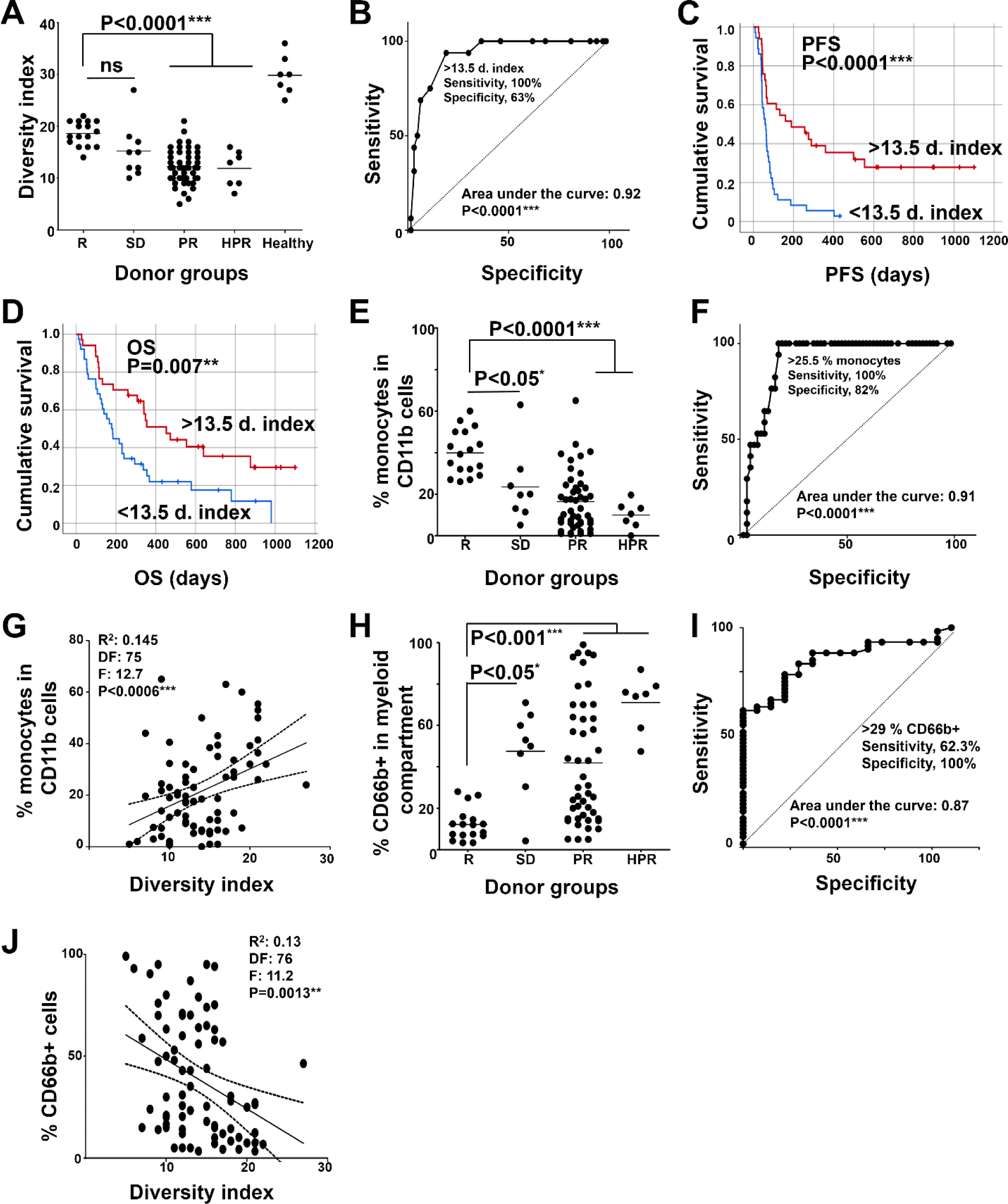
Baseline diversity of myeloid cells in NSCLC patients and correlation with clinical responses. (**A**) Diversity indexes as a function of clinical responses. **(B)** ROC curve of the diversity index as a function of objective responses vs non-objective responses. **(C)** Kaplan-Meier plot of PFS stratifying the patients according to high or low diversity index. n=69 (33 above the cut-off, 36 below). **(D)** As in (C) but plotting OS. N= 72 (34 above the cut-off, 38 below). **(E)** Percentage of circulating monocytes (CD11b^+^ CD14^+^ HLA-DR^+^) within each response group as indicated. **(F)** ROC analysis of the percentage of monocytes as a predictor of objective responses. **(G)** Correlation between the percentage of monocytes and diversity index. **(H)** Percentage of granulocytic cells (CD11b^+^ CD66b^+^) within each response group as indicated. **(I)** ROC analysis of the percentage of granulocytic myeloid cells as a predictor of no objective response. **(J)** Correlation between the percentage of granulocytic myeloid cells and diversity index as a predictor of no objective response. R, objective responders (n=16); SD, stable disease (n=9); PR, progressors (n=47); HPR, hyperprogressors (n=7); Healthy, represents a sample of age-matched donors without known pathologies (n=7); ns, non-statistical differences. Relevant statistical comparisons are shown within the graphs. Multicomparisons in dot-plots were carried out by the Wilcoxon test. Pair-wise comparisons were performed by Mann-Whitney’s U test. Survival differences were tested with the Log-rank test, and correlation plots with Spearman’s test. *, **, ***, indicate significant (p<0.05), very significant (p<0.01) and highly significant (p<0.001) differences.

Objective responders had significantly (p<0.0001) elevated baseline percentages of monocytes within CD11b^+^ cells (39.86% ± 10.85; Mean; Mean ± SD) compared to patients with stable disease, progressors (16.41% ± 14.14;; Mean ± SD) and hyperprogressors. In agreement with other studies (*17, 40*), it had predictive value in our discovery cohort (**Fig. 1E**, **Fig. S3**). A cut-off value of >25% of monocytes identified responders with 100% sensitivity and 82% specificity, according to ROC analysis (AUC=0.9069; 95% confidence interval = 0.8424 to 0.9715; p<0.0001) (**Fig. 1F**). To test whether increased monocytic cells was characteristic of elevated systemic myeloid diversity, the percentage of CD14^+^ HLA-DR^+^ monocytes within CD11b^+^ cells was correlated to diversity indexes, demonstrating a significant (p<0.0006) positive correlation (**Fig. 1G**). On the other hand, patients with stable disease, progressors and hyperprogressors presented more than 50% of circulating CD66b^+^ cells, corresponding to neutrophils and granulocytic MDSCs, compared to an average 12.20% ± 7.76 (Mean ± SD) in responders, which represent a significantly (p<0.001) higher frequency of circulating CD66b^+^ cells in non-responders (**Fig 1H**, **Fig. S3**). A cut-off value of >29% of CD66b^+^ cells identified non-responders with 100% sensitivity and 62% specificity, according to ROC analysis (AUC=0.8689; 95% confidence interval 0.7878 to 0.9499; p<0.0001) (**Fig. 1I**). Likewise, the DI was also significantly (p=0.0013) associated to reduced percentages of systemic CD66b^+^ granulocytic cells (**Fig. 1J**). These data confirmed that objective responders showed a systemic myeloid signature dominated by monocytic CD14^+^ HLA-DR^+^ cells accompanied with low abundance of granulocytic CD66b^+^ myeloid cells (neutrophils and G-MDSCs), while patients with stable disease, progressors and hyperprogressors showed differential myeloid profiles **(Fig. S3)**.

### Plasma FKN concentration is associated with myeloid diversity and objective responses

To identify profiles of plasma soluble factors that could differentially impact over myeloid diversity and functions in patients, a panel of 65 cytokines, chemokines and soluble immune checkpoints was evaluated in plasma before the start of immunotherapy, and after the first cycle of therapy. While most of the quantified plasma soluble analytes showed increased levels in patients who rendered no objective clinical responses to anti PD-1/PD-L1 immunotherapy, some of them, such as GM-CSF, G-CSF and IL12p70, were elevated in a sample of responders before treatment, but only FKN concentrations reached statistical significance (p=0.0037) **(Fig. 2A) (Table S2)**. FKN plasma levels in clinical samples positively correlated with myeloid diversity in our discovery cohort (**Fig. 2B**). A cut-off value for FKN concentration above 55 pg/ml was identified by ROC analysis (**Fig. 2C**) for significant benefit in PFS (p=0.003) and OS (p<0.001) (**Fig. 2D, E**). Our cohort was recruited through an exploratory prospective study that inherently had variability in the patient cohort. Therefore, to validate our results FKN transcriptomic expression was analyzed in lung cancer clinical samples registered in The Cancer Genome Atlas (TCGA) database. We found that high FKN mRNA levels were significantly (p=0.0132) associated to prolonged survival rates in a cohort of 515 lung adenocarcinoma (LUAD) patients. This cohort included different stages and treatment protocols, thus supporting our data and but also highlighting the prognostic value of FKN transcription independently of the treatment (**Fig. 2F**). Differences were not found in a sample of squamous lung carcinoma (LUSC) patients (**Fig. 2G**). Furthermore, we identified 8 additional biomarkers that significantly differed between patients who rendered objective partial responses to immunotherapy from those who showed tumor progression **(Fig. S4)**. Most of the other analyzed analytes did not discriminate between these two groups **(Table S2)**.

**Figure 2.**
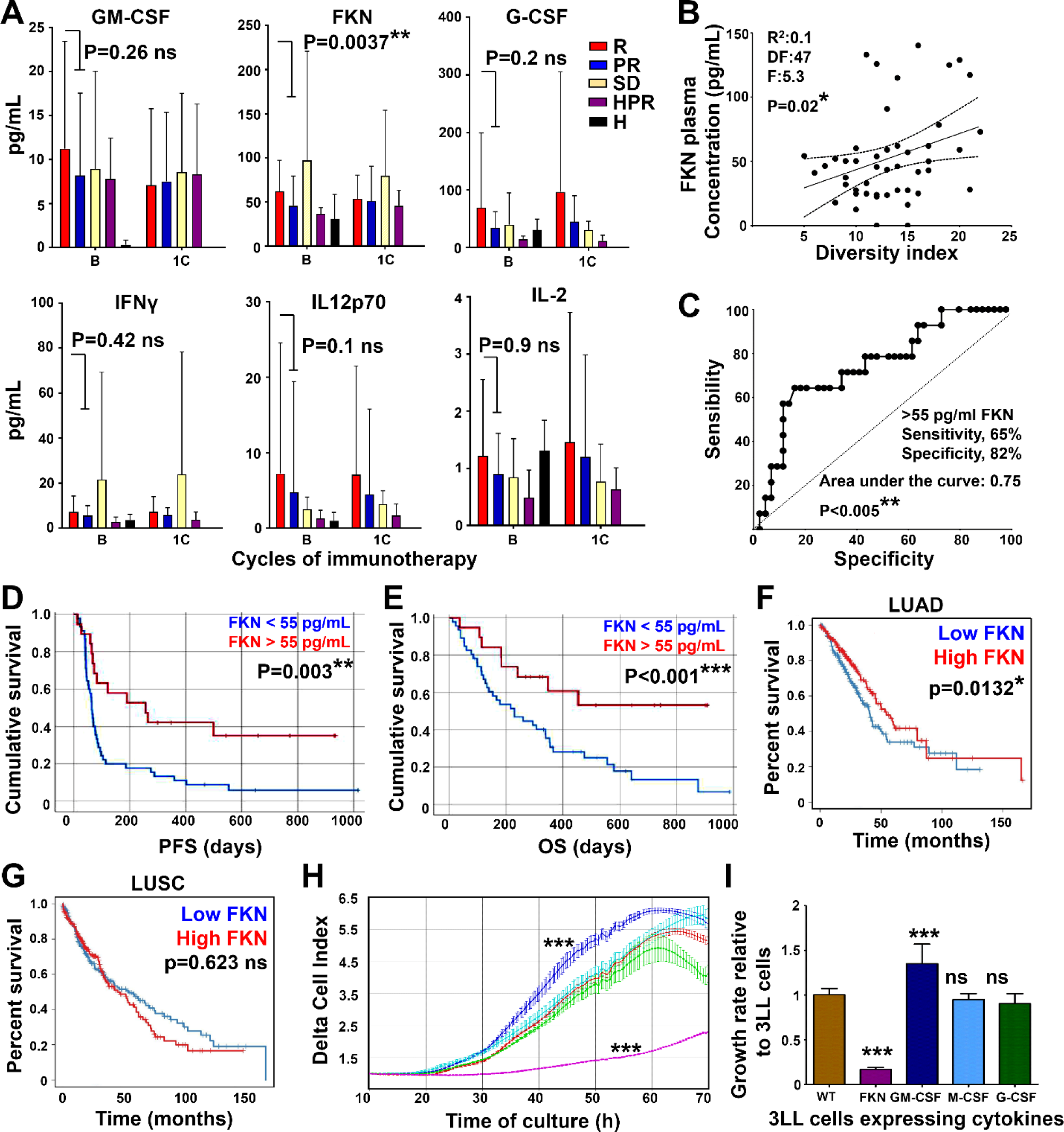
Quantification of systemic plasma factors with myeloid-regulating activities in NSCLC patient cohorts. **(A)** The bar graphs show plasma concentrations of the selected proteins in responders (R, n=17, 26), progressors (PR, n=38, 59), hyperprogressors (HPR, n=3), patients with stable disease (SD, n=5, 11) and age-matched healthy donors (H, n=4, 6) before (baseline, B) and after the first cycle of immunotherapy (1C). Error bars are shown (standard deviations, SD), and relevant statistical comparisons by Mann-Whitney U tests are shown within the graphs. **(B)** Correlation between FKN plasma concentration and diversity index with Spearman’s test. **(C)** ROC analysis of FKN concentration as a predictor of objective responses. The calculated cut-off value for the statistics provided in the graph is shown. **(D)** Kaplan-Meier plot of PFS stratifying the patients according to the FKN cut-off value identified in the ROC curve (N=42). The associated P value is shown. **(E)** As in (D) but plotting OS. n= 46 (17 above the cut-off, 29 below). **(F)** Kaplan-Meier survival plot of lung adenocarcinoma (LUAD) patients. Patients were stratified according to FKN transcriptional expression level within tumor samples. Data were retrieved from the TCGA database and exported with TIMER2.0. Differences in survival were evaluated with the Log rank test, and associated P values are shown within the graph. n=515. HR=0.827; 95% confidence interval=0.784 to 0.945. **(G)** As in (F) but plotting squamous lung carcinoma (LUSC) patients n=501. HR=1.03; 95% confidence interval=0.885 to 1.095. **(H)** Real time cell growth (RTCA) of 3LL cell lines engineered to secrete the indicated cytokines. Relevant statistical comparisons of delta-cell indexes after 50 h of culture were carried out by ANOVA. **(I)** Bar graphs with cell growth rates for the indicated 3LL cell lines relative to the growth of unmodified 3LL cells. Error bars are shown (SD). Statistical comparisons by ANOVA and Tukey’s pairwise comparison tests are indicated. *, **, ***, indicate significant (p<0.05), very significant (p<0.01) and highly significant (p<0.001) differences; ns, non-significant differences.

Since a tendency for increased levels of hematopoietic growth factors such as GM-CSF and G-CSF were identified in the plasma of responder patients, and a correlation between FKN levels and myeloid diversity was also found, the effects of FKN were tested on cancer cells through overexpression. As controls, cancer cell lines overexpressing hematopoietic growth factors were also generated. To that end, murine adenocarcinoma 3LL cells were engineered to constitutively overexpress GM-CSF, G-CSF, M-CSF or FKN **(Fig. S5)**. We found that only GM-CSF and FKN expression significantly but inversely altered the *in vitro* growth rate of 3LL cells (**Fig. 2H**). While FKN was a very strong inhibitor of cell growth, GM-CSF was a significant enhancer of 3LL proliferation (**Fig. 2I**).

### FKN impairs lung cancer cell growth and increases immune cell diversity *in vivo*

We investigated the potential anti-oncogenic mechanisms of FKN in lung cancer and its use as a therapeutic agent. We first demonstrated that targeting the CX3CL1-CX3CR1 signaling axis with recombinant FKN in a murine model of lung adenocarcinoma altered *in vivo* tumor growth. A single dose of recombinant FKN was sufficient to delay 3LL tumor growth. In contrast, the pharmacological inhibition of CX3CR1 by the administration of the allosteric non-competitive FKN antagonist AZD8797 accelerated tumor growth (p<0.05) (**Fig. 3A**). Then, 3LL cell lines expressing FKN, GM-CSF, G-CSF and M-CSF were subcutaneously transplanted in groups of mice. FKN secretion strongly impaired (p=0.0002) 3LL tumor growth leading to increased median survival from 17 days in the control group to 30.5 days in the FKN overexpressing group (p=0.0005) (**Fig. 3B, C**). G-CSF and M-CSF expression also delayed tumor growth and increased survival, but to lesser extent. GM-CSF expression by 3LL cells significantly accelerated tumor progression. These results demonstrated that FKN had potent anti-tumor activities both directly as a recombinant protein, and through expression by tumor cells. FKN-producing tumor-bearing mice showed elevated concentrations (0.85 ng/mL ± 0.11) of plasma soluble FKN compared to mice engrafted with unmodified 3LL cells (0.36 ng/mL ± 0.05), and not higher than that from naive mice (1.32 ng/mL ± 0.03) (**Fig. 3D**), as expected due to the small size of tumors at the selected time point (**Fig. 3E**). Other plasma soluble cytokines showed no significant differences between FKN-producing tumor-bearing mice and their wild-type counterparts **(Fig. S6).** These results confirmed FKN anti-tumor activities both systemically as a recombinant protein but also through local production within the tumor microenvironment.

**Figure 3.**
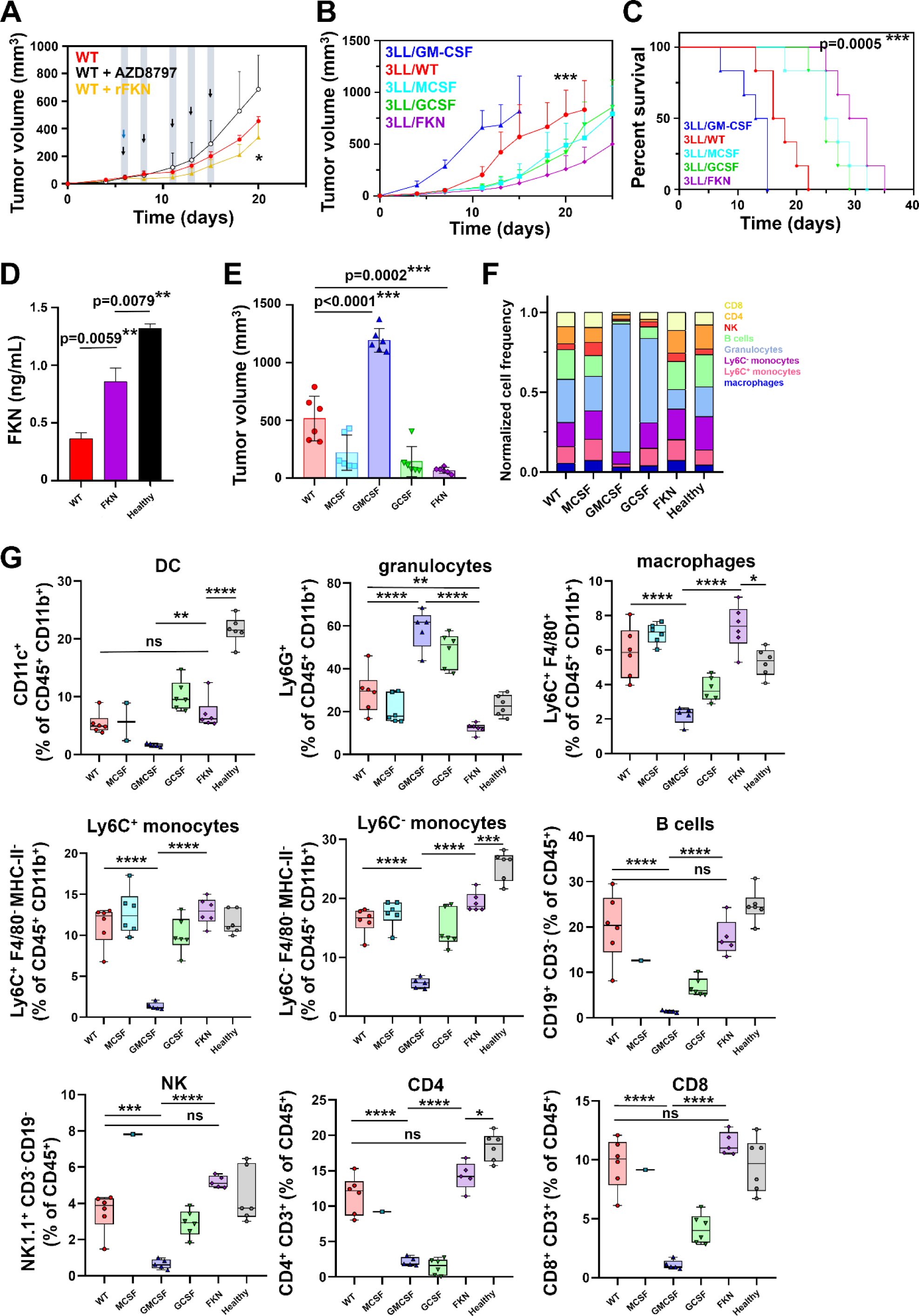
FKN inhibits in vivo lung cancer cell growth, characterized by an increase of systemic immune cell diversity and a specific cytokine profile. (**A**) Tumor growth curves of 3LL-tumor bearing mice treated with recombinant FKN (rFKN) or with the CX3CR1 inhibitor AZD8797. Arrows indicate the time of intraperitoneal injections of rFKN (blue) and AZD8797 (black). Data are expressed as means ± SD (n=6 mice per group). Statistical comparisons by ANOVA followed by Tukey’s pairwise comparison tests are provided. (**B**) In vivo tumor growth of engrafted 3LL cell lines producing FKN and key myelopoiesis-driving cytokines as indicated. Data are expressed as mean ± SD (n=6 mice per group). Comparisons between groups were performed by ANOVA and Tukey’s pairwise comparison tests. (**C**) Kaplan-Meier survival plots. Differences between the control group (WT) and the FKN group were evaluated by a two-sided log-rank test. Statistical comparisons are shown within the graph. (**D**) FKN plasma concentration at day 15 in the indicated groups of tumor-bearing mice. Data are expressed as mean ± SD from a pool of 6 mice/group. Statistical comparisons were performed by ANOVA and Tukey’s pairwise comparison tests. (**E**) Tumor volumes 15 days after tumor inoculation. Data are expressed as mean ± SD (n=6 mice per group), and comparisons between groups were performed by ANOVA and Tukey’s pairwise comparison tests. (**F**) Myeloid and lymphoid diversity in peripheral blood at day 15 in mice transplanted with 3LL-expressing the indicated cytokines. WT, unmodified 3LL cells; Healthy, mice without tumors. (**G**) Box and whisker plots of the percentage of the indicated peripheral immune cell populations at day 15 after injection of groups of mice (n=6 mice per group) with 3LL cells overexpressing the indicating myeloid-regulating cytokines. The relevant immune populations were quantified as percentages of total leukocytes (CD45^+^ cells) into DCs (CD11c), macrophages (F4/80), Ly6C^+^ monocytes and Ly6C^-^ monocytes, granulocytes (Ly6G), B cells (CD19), NKs (NK1.1), CD4 T cells (CD3 CD4) and CD8 T cells (CD3 CD8). Relevant statistical comparisons were performed by Wilcoxon’s test followed by pair-wise comparisons of relevance by Mann-Whitney U test. *, **, ***, ****, indicate significant (p<0.05), very significant (p<0.01), highly significant (p<0.001) and very highly significant (p<0.0001) differences; ns, non-significant differences.

Profiling of circulating immune cells was performed when significant differences in tumor volume were observed (**Fig. 3E**). Similar to healthy controls, FKN-producing tumor-bearing mice presented balanced phenotypic profiles within myeloid and non-myeloid cell compartments (**Fig. 3F**). These results were also consistent with the increased cell diversity found in responder patients within our discovery cohort. In fact, 3LL-FKN tumor bearing mice showed decreased Ly6G^+^ granulocytes (12.25% ± 2.36) compared to their wild-type counterparts (29.2% ± 9.9) (**Fig. 3G**), whereas 3LL/GM-CSF tumor bearing mice exhibited a landscape consistent with human hyperprogressors, dominated by an exponentially accelerated (p<0.0001) tumor growth, shortened survival (median survival of 14 days vs 17 days of wild type control, p=0.009) and overwhelming numbers of MDSCs (58.52% ± 9.17 of Ly6G^+^ cells).

### FKN synergizes with anti-PD1 immunotherapy

Our clinical data associated plasma FKN concentrations superior than 55 pg/mL with objective responses to PD-L1/PD-1 blockade. Therefore, we investigated whether FKN contributed to PD-L1/PD-1 blockade efficacy in the murine 3LL lung adenocarcinoma model. FKN expression by cancer cells caused an evident delay (p<0.0001) in tumor growth with a significant (p=0.0014) increase in median survival from 20 to 35 days. Importantly, while naïve tumors were refractory to PD-1 monoblockade *in vivo*, FKN sensitized 3LL tumors to PD-1 blockade (**Fig. 4A, B**). This effect was not observed for soluble forms of GM-CSF, G-CSF and M-CSF **(Fig. S7)**.

**Figure 4.**
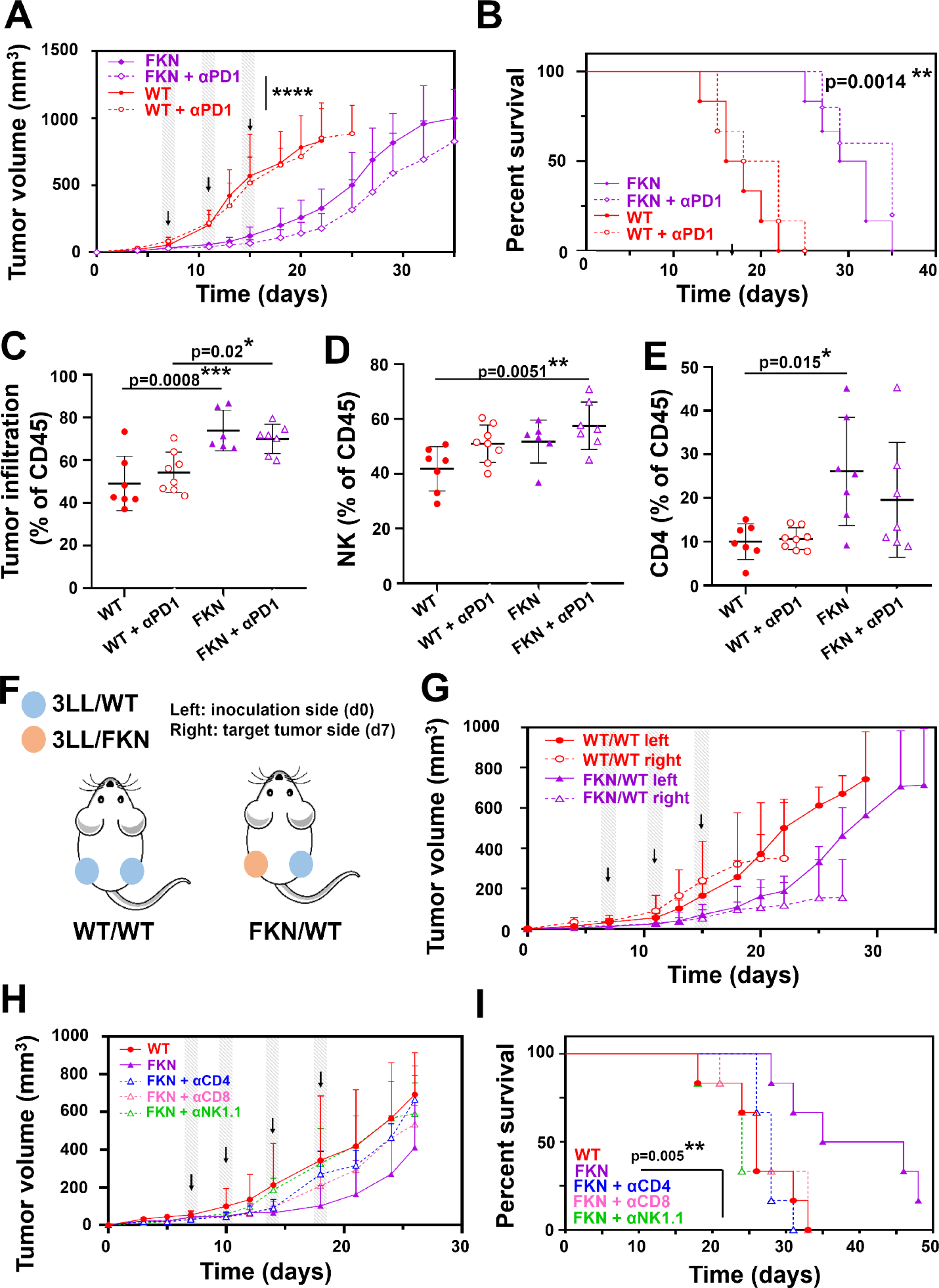
In vivo anti-tumor mechanisms of FKN in combination with PD-1 blockade in a murine model of lung adenocarcinoma. **(A)** Unmodified and FKN-expressing 3LL cells were subcutaneously inoculated in mice and tumors were allowed to grow for 7 days. Tumor-engrafted mice were treated intraperitoneally with anti-PD-1 antibody or vehicle (days 7, 11 and 15, as indicated by arrows) following randomization into two groups. Tumor growth was monitored. Data are presented as mean ± SD (n=6 mice per group). **(B)** Kaplan-Meier survival plots of the indicated groups of mice. Survival differences between the control group (WT + αPD-1) and FKN + αPD-1 were evaluated by a two-sided log-rank test. **(C)** Percentage of tumor infiltration with total leukocytes (CD45^+^), **(D)** NK cells (NK1.1^+^) and **(E)** CD4 T lymphocytes. Infiltration data are shown as mean ± SD (n=8 mice per group). Relevant statistical comparisons are shown within the graphs with ANOVA and Tukey’s pairwise comparisons. **(F)** Experimental schedule for testing abscopal anti-tumor activities. 3LL-parental (WT) or 3LL- FKN (FKN) cells were subcutaneously injected on the left flank of mice (inoculation side). 7 days after later, 3LL-parental cells were engrafted on the right flank (target tumor side). Mice were intraperitoneally treated with anti-PD-1 antibody at days 7, 11 and 15 after the last tumor inoculation. **(G)** Tumor growth of right and left flank engrafted tumors in the experimental schedule shown in (f). Data are presented as mean ± SD (n=6 mice per group). **(H)** Tumor growth in mice intraperitoneally treated with anti-CD4, anti-CD8 and anti-NK1.1 depleting antibodies at days 6, 10, 14 and 18 after tumor inoculation, as indicated by arrows. Data are presented as means ± SD (n=6 mice per group). **(I)** Kaplan-Meier survival plot of the indicated treatment groups. Survival differences between the control group (WT) and FKN were evaluated by two-sided log-rank test. *, **, ***, indicate significant (p<0.05), very significant (p<0.01) and highly significant (p<0.001) differences; ns, non-significant differences.

FKN is known to attract immune cells to the tumor microenvironment^30^. Therefore, the tumor immune infiltrate was characterized at a time point with significant differences in tumor volumes. Only mice engrafted with FKN-producing cells showed increased immune cell infiltration within tumors especially with NK and CD4 T cells (**Fig. 4C-E** **and Fig. S8).** Our data suggested that plasma soluble FKN acts systemically to potentiate the antitumor activity of PD- L1/PD-1 blockade. To test this hypothesis, 3LL-FKN cells were subcutaneously implanted in mice on the left flank (inoculation side), followed by injection of unmodified 3LL cells on the right flank (target tumor side) one week later (**Fig. 4F**). Mice were then treated with anti-PD-1 immunotherapy and tumor growth was monitored at both flanks. Interestingly, the growth of 3LL tumors was delayed, and comparable to 3LL-FKN tumors in the same mice (**Fig. 4G**). These results demonstrated abscopal effects of FKN associated with anti-PD-1 blockade. Hence, FKN acts in synergy with PD-1 blockade systemically, as well as promoting immune infiltration of tumors with NK cells.

Previous studies implicate several immune cell types in FKN-mediated anti-tumor mechanisms such as CX3CR1^+^ T cells, DCs and NKs (*22-25, 28-30, 41-43*). To identify the anti-tumor effector cells in our experimental systems, CD4, CD8 and NK cells were depleted in mice and the growth of 3LL-FKN tumors was monitored. Interestingly, NK cell abrogation completely restored the growth of 3LL-FKN tumors to rates comparable to unmodified 3LL tumors (**Fig. 4H**). Limited effects were observed by CD4 and CD8 T cell depletion. These results suggested that the *in vivo* inhibition of tumor growth by secreted FKN was mainly caused by NK activity, with a minor contribution by T cells. Our results were consistent with NK cells as prominent systemic mediators of FKN-driven anti-tumor immunity. Indeed, NK depletion shortened median mice survival down to 24 days, compared to 40.5 days of median survival in the FKN control group (p=0.0051) (**Fig. 4I**).

To validate our results of FKN-driven tumor infiltration in mice, we analyzed transcriptional data from lung adenocarcinoma samples included in the TCGA database. Correlation analyses of FKN transcriptomic expression with immune infiltrates was performed with the Tumor Immune Estimation Resource tool (TIMER2.0) (*44*). A significant positive association of FKN expression with CD8 (Rho = 0.222, p=6.28e-07), CD4 (Rho=0.185, p=3.66e-05), NK (Rho=0.244, p=4.07e-08), neutrophil (Rho=0.211, p=2.39e-06) and monocyte infiltration (Rho=0.263, p=2.96e-09) was confirmed. In contrast, a negative correlation with MDSC infiltration was observed (Rho=- 0.269, p=1.21e-09). Infiltration with T_reg_, B lymphocytes, macrophages and myeloid DCs populations was also analyzed with differential results (**Fig. 5A**).

**Figure 5.**
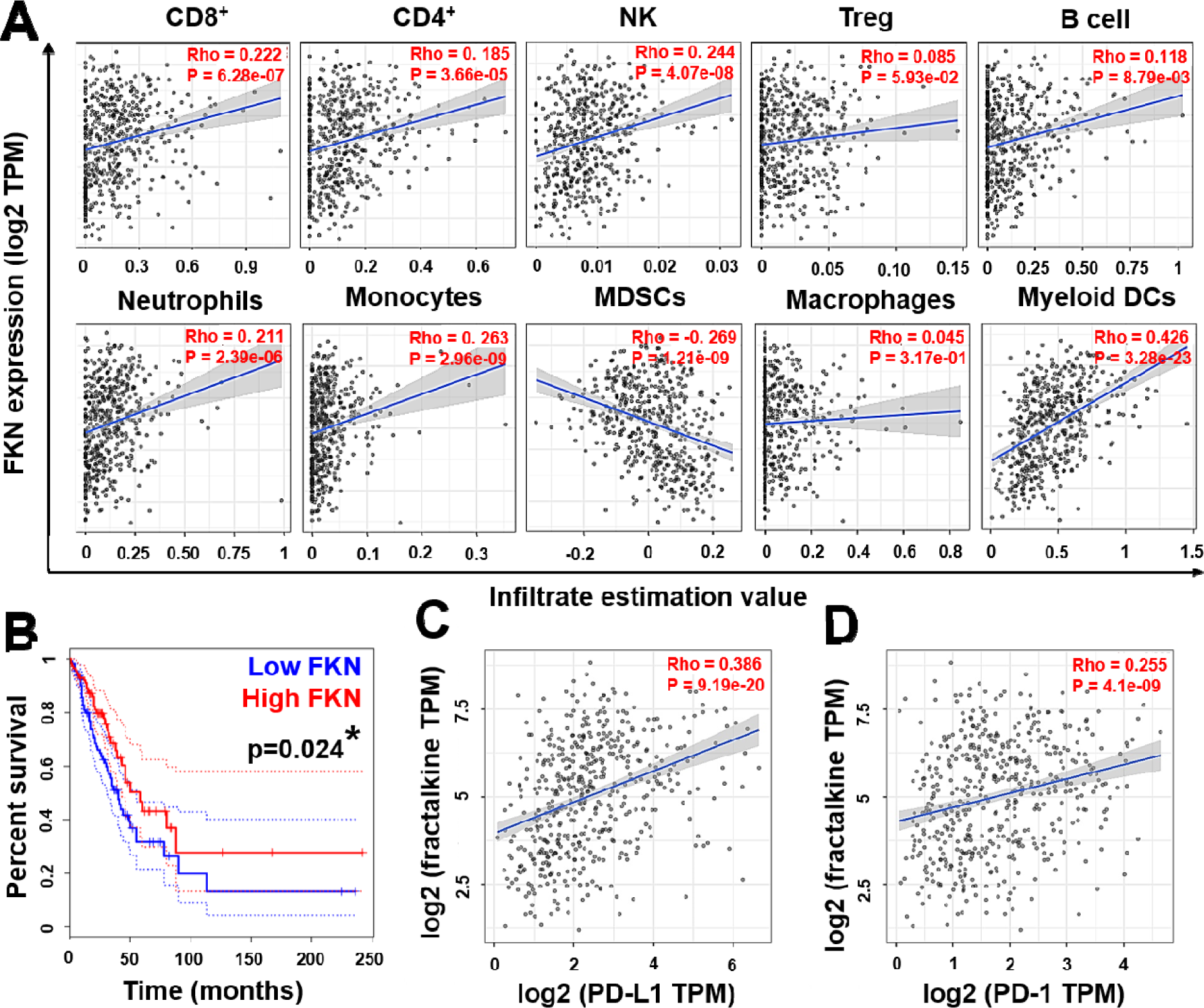
FKN transcriptomic expression in human lung adenocarcinoma samples and correlation with tumor immune infiltration, survival and PD-L1 tumor expression. (**A**) Evaluation of tumor infiltration with the indicated selected immune cell populations, and correlation analyses of FKN transcriptional expression and immune infiltrates from the TCGA database. Analyses were restricted to lung adenocarcinoma samples (n=515). Spearman correlation with different immune populations were identified by several algorithms (CIBERSORT, quanTIseq, xCell, TIDE) and an adjustment based on tumor purity was employed to minimize the potential interaction of low tumor cell quantities. Each dot represents a single tumor sample. Spearman’s rho value and p values are provided within the graphs. (**B**) Kaplan-Meier plot of OS according to FKN expression in a lung adenocarcinoma cohort registered in the TCGA database (n=240). Patients were stratified according to the median FKN expression value. Survival differences were tested by the Log rank test. Dotted lines represent 95% confidence intervals. (**C**) Correlation between tumor FKN and PD-L1 transcriptional expression by Spearman’s test. Relevant statistical results are presented within the graphs. (**D**) As in (C) but plotting PD-1 transcriptional expression levels.

We also correlated the degree of FKN transcriptomic expression in human tumor samples with survival with the Gene Expression Profiling Interactive Analysis tool (GEPIA2) (*45*). In agreement with our findings, high median FKN mRNA expression levels in tumors were associated with better OS (**Fig. 5B**). PD-L1 tumor expresssion is a clinical biomarker used to predict response to PD-L1/PD-1 blockade (*46*). Importantly, FKN and PD-L1 expression followed a positive correlation in tumor samples (**Fig. 5C**) as did PD-1 (**Fig. 5D**), supporting our results in the discovery cohort and the potential synergy of FKN expression with the PD-1/PD- L1 blockade axis.

### FKN plasma concentration as a biomarker in lung adenocarcinoma compared to squamous lung cancer patients treated with PD-L1/PD-1 blockade

Recent reports highlight that stratification of histological lung cancer sub-types have to be taken into consideration in discovery studies of novel biomarkers (*47*). Classically, lung squamous and lung adenocarcinomas are grouped together under the term NSCLC. Nevertheless, since LUSC is a less frequent malignancy among lung cancer patients, this classification could be masking the value of biomarkers aimed to NSCLC. Indeed, there were differences in survival in our cohort depending on tumor histology (**Fig. 2F, G**). Therefore, to evaluate whether plasma FKN concentration as a biomarker of response was dependent on tumor histology in NSCLC, our discovery cohort was stratified in two groups, adenocarcinoma and squamous cell lung cancer. As expected, longer PFS and OS correlated with FKN plasma concentrations above the previously established cut-off value (55 pg/mL) in lung adenocarcinoma patients (**Fig. 2D, E**). No significant correlation was observed in lung squamous cell carcinoma patients (**Fig. 6A, B**). These results were also consistent with previous studies (*48*). Next, we tested whether *in vitro* FKN production by cancer cell lines was dependent on histology. FKN secreted by 5 LUAD and 5 LUSC human cancer cell lines was evaluated by ELISA. Soluble FKN production was cell line-dependent although LUAD cell lines tended to secrete higher FKN levels than LUSC cell lines (**Fig. 6C**).

**Figure 6.**
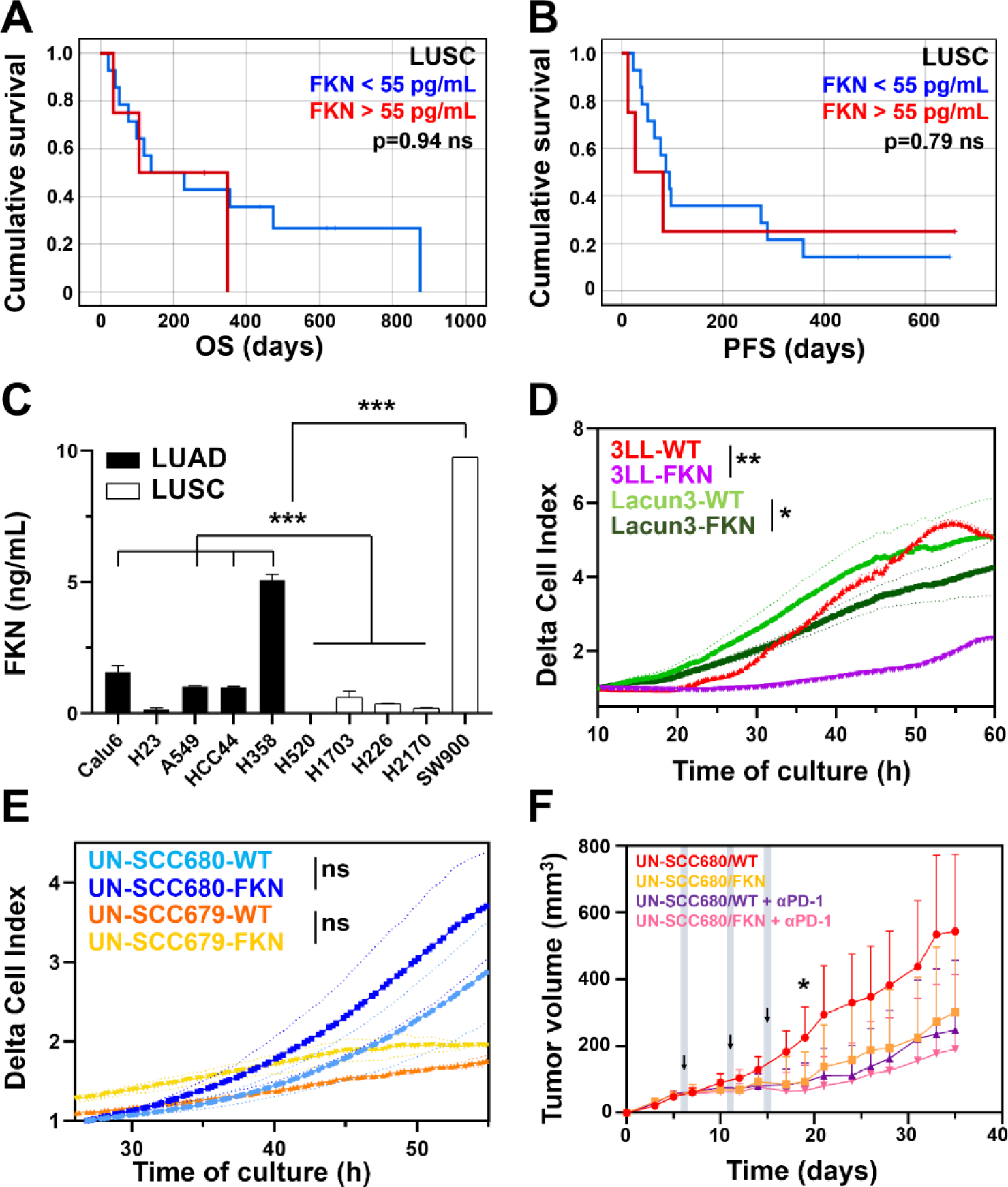
lasma fractalkine concentration as biomarker of response in different lung cancer histologies. **(A)** OS plots of our discovery cohort of LUSC patients stratified according to baseline FKN plasma concentrations. n=18 (11 below median, 7 above). **(B)** As in (A) but plotting PFS. **(C)** Quantification of secreted FKN in cell cultures of a collection of 10 LUAD and LUSC cancer cell lines, quantified by ELISA. Results are presented as means ± SD. Samples were assayed in duplicates from a pool of three independent replicates. Statistical differences among groups were analyzed by ANOVA followed by Tukey’s tests. **(D)** Proliferation of two LUAD murine cell lines overexpressing FKN and their parental unmodified counterparts (WT), as indicated. Real-time cell growth was monitored by RTCA. Delta cell indexes are expressed as mean ± SD from three independent replicates. Differences in delta cell index proliferation data were tested by ANOVA following Tukey’s pairwise comparison tests after 40 h of culture. **(E)** Proliferation of two LUSC murine cell lines overexpressing FKN and their parental unmodified counterparts (WT), as indicated. Real-time cell growth was monitored by RTCA. Delta cell indexes are expressed as mean ± SD from three independent replicates. Differences in delta cell index proliferation data was tested by ANOVA following Tukey’s pairwise comparison tests after 50 h of culture. **(F)** Tumor growth from parental and FKN-expressing UN-SCC680 cells inoculated in A/J mice. Tumors were allowed to grow for 6 days, and then mice were randomized into two groups. After randomization, mice were treated with anti-PD-1 antibody or vehicle (days 6, 11 and 15). Data are presented as mean ± SD (n=6 mice per group). Differences in tumor growth were evaluated by ANOVA and Tukey’s pairwise comparisons at day 19 after tumor inoculation. Relevant statistical comparisons are shown within the graphs. *, **, ***, indicate significant (p<0.05), very significant (p<0.01) and highly significant (p<0.001) differences; ns, non-significant differences.

To further elucidate the effect of FKN in tumors with different histological origins, we engineered the murine adenocarcinoma cell line Lacun3 (*49*) and two squamous lung cancer cell lines, UN-SCC679 and UN-SCC680 (*50*), to constitutively overexpress FKN **(Fig. S5)**. We confirmed that FKN had differential effects over the *in vitro* growth of cancer cells depending on the histology, consistent with our clinical data. The two FKN-expressing LUAD cell lines showed impaired cell growth *in vitro* (**Fig. 6D**), while LUSC cell lines expressing FKN exhibited increased growth rates (**Fig. 6E**). Furthermore, LUSC UN-SCC680 cells showed increased invasion potential *in vitro* compared to 3LL LUAD cells **(Fig. S9)**. Surprisingly, FKN expression did not accelerate tumor growth *in vivo* in LUSC UN-SCC680-tumor bearing mice with or without PD-1 blockade (**Fig. 6F**). These results suggested that efficacious anti-tumor immune responses may still take place in a tumor histology-independent manner. All together, these results suggested that even though plasma soluble FKN may not have value as a biomarker of response in LUSC patients for PD-L1/PD-1 blockade immunotherapies, a therapeutic role for this cytokine could not be discarded. UN-SCC680-derived tumors were highly sensitive to PD-1 blockade monotherapies, as previously reported (*50*). Hence, only moderate synergy with FKN expression was observed (**Fig. 6F**).

## DISCUSSION

Understanding the mechanisms underlying clinical failure and adverse events in PD-L1/PD-1 blockade immunotherapies represents a major challenge. Increasing evidence indicates that functional systemic immunity is required for the success of immunotherapies (*1, 2, 4, 6-11*). For example, dysfunctional peripheral blood CD4 and CD8 T cells prior to immunotherapies are biomarkers of clinical failure in PD-L1/PD-1 monotherapies, as shown by us and others (*4, 6, 7, 10*). Likewise, systemic expansion of myeloid suppressor cells and neutrophils constitute poor prognostic markers not restricted to immunotherapies (*51-53*). MDSCs and granulocytes are strong T cell suppressors in cancer through several mechanisms, as extensively reviewed elsewhere (*54, 55*). On the other hand, elevated percentages of HLA-DR^+^ monocytes correlate with efficacious PD-1 blockade immunotherapy (*17*). These monocytic subsets are proficient T cell activators through strong antigen-presenting capacities. Hence, profiling of immune cell composition and functionality in peripheral blood could be used for selection of potential responder patients in PD-L1/PD-1 blockade immunotherapies. Following this reasoning, we performed detailed analyses of myeloid cell subsets and their differentiation stages in a discovery cohort of NSCLC patients before starting immunotherapies by high-dimensional flow cytometry. A DI was derived to reflect the variety of phenotypes and activation stages by high-dimensional cluster analyses in lung cancer patients and healthy donors. Here we confirmed that an elevated diversity of circulating myeloid cell types was characteristic of objective responders to PD- L1/PD-1 blockade. It could be argued that a diverse composition of peripheral immune cell phenotypes could reflect functional myelopoiesis (*56*) in patients before starting ICB. Indeed, elevated diversity of phenotypes was particularly represented by an enrichment of activated classical monocytes with a concomitant decrease in granulocytic myeloid cells. Our data confirmed that in addition to their classical prognostic value, these myeloid cell types show significant predictive power in PD-L1/PD-1 blockade immunotherapies by ROC analyses, in agreement with previous studies with high-dimensional analytical techniques (*17, 57, 58*). Our high-dimensional profiles included (but were not restricted to) DC, monocytes, macrophages, neutrophils and MDSCs at various differentiation and activation stages, and demonstrated that the balance between these immune cell types is critical for ICB outcome before starting immunotherapies.

On the lookout for plasma factors associated with (1) high systemic myeloid cell diversity, (2) elevated monocyte populations and (3) increased survival, we found that only FKN fulfilled these three conditions. Hence, in our discovery cohort FKN plasma concentration showed good predictive value for PD-L1/PD-1 blockade therapies in NSCLC patients as shown by ROC analyses. Moreover, this result was independently corroborated by correlates of FKN transcription with survival and tumor PD-L1 expression obtained from the TCGA database, amounting to 515 human lung adenocarcinoma samples. In agreement with the clinical data, mice engrafted with FKN-producing tumors showed more balanced myeloid and lymphoid circulating cell subsets, with a systemic drop in Ly6G^+^ granulocytes and increased OS.

FKN showed strong anti-cancer properties both *in vitro* and in *in vivo* models for murine lung adenocarcinoma and squamous cell lung cancer. Adenocarcinoma 3LL cells overexpressing FKN showed reduced growth *in vitro* and *in vivo*, while FKN expression by UN-SCC680 squamous carcinoma cells impaired tumor growth *in vivo*. These results were not caused by exogenous overexpression or by the genetic modification of cancer cells, as systemic injection of recombinant FKN directly delayed 3LL tumors in mice. In agreement with this result, systemic administration of a FKN receptor inhibitor counteracted the inhibitory effects of soluble FKN over the growth of adenocarcinoma tumors.

Our study demonstrated that FKN possessed systemic anti-tumor activities, without the need to be expressed within the tumor microenvironment. This is important, because it was previously thought that FKN expressed by tumors could exert anti-tumor effects through recruitment of T cells, DCs and NK cells to the tumor microenvironment (*22-30*). In addition to its systemic effects, we also observed increased tumor infiltration especially by NK cells and CD4 T cells. Our data in mouse models were corroborated by correlation analyses of FKN transcriptomic expression with immune infiltrates. Nevertheless, we demonstrated that anti-tumor systemic and abscopal effects were caused mainly by circulating NK cells, with a minor contribution by CD4 and CD8 T cells. It has been shown in some tumor types that the membrane-bound FKN can promote metastasis of CX3CR1^+^ circulating tumor cells towards tissues displaying a high CX3CL1 expression, such as bones, lungs and nervous tissues (*31-38*). Our data clearly showed that at least in LUAD and LUSC cancer models this was not the case. This dual behavior of FKN may probably be a consequence of the two different bioactive forms of the protein (*59-61*). The membrane-bound form mediates cell adhesion processes and migration while the soluble version participates in chemoattraction of effector cells to the tumor, but also in activating systemic NK cell responses as shown here.

Our clinical data showed that plasma FKN concentration had good predictive value in NSCLC treated with ICB as shown by ROC analyses. The most frequent histological subtype of lung cancer corresponds to LUAD, while the minority correspond to LUSC. Recent studies are shedding light into the numerous differences between them that would require the identification of biomarkers of response with discriminating values for each histology independently (*47, 62, 63*). Indeed, plasma FKN had positive predictive value only for LUAD treated with ICB in our discovery cohort. We also confirmed that FKN transcription in lung cancer tumor samples from

TCGA was associated to better survival. These data agreed with the only transcriptomic study reporting a differential value of FKN tumor expression as a prognostic biomarker for LUAD and LUSC (*48*).

Importantly, we demonstrated that secreted FKN rendered 3LL LUAD-derived tumors sensitive to PD-1 blockade, a model highly refractory to PD-L1/PD-1 monotherapies (*64*). We observed similar results for UN-SCC680 LUSC-derived tumors, although in this case PD-1 blockade had significant therapeutic effects on its own (*50*). We concluded that although FKN plasma levels had differing predictive value in LUAD and LUSC in human patients, the therapeutic use of FKN was not altered by tumor histology in mouse lung cancer models.

Concluding, plasma FKN is a biomarker of systemic myeloid cell diversity with elevated activated monocytic cell subsets and decreased MDSCs. It correlates with increased survival in human lung adenocarcinoma patients treated with PD-L1/PD-1 blockade monotherapies. Our results demonstrate a significant therapeutic effect of soluble FKN not restricted to the tumor microenvironment, mainly through the activity of NK cells. These results open an avenue towards the use of plasma FKN concentration as a biomarker of response, and its therapeutic use in combination with anti-PD-1/PD-L1 immunotherapies in NSCLC patients. The results obtained here were obtained from a prospective observation study in a discovery cohort, and supported through analyses of transcriptomic data. Nevertheless, the use of FKN as a biomarker of response will require proper validation in a randomized multicentre clinical trial.

## MATERIALS AND METHODS

### Clinical samples and study design

One hundred and twelve patients diagnosed with locally advanced or metastatic NSCLC treated with immune checkpoint inhibitors (ICI) nivolumab, pembrolizumab and atezolizumab, or with the combination of chemo-immunotherapy (pembrolizumab + platinum-based chemotherapy) were recruited between December 2017 and February 2021 at the Oncology Department of the University Hospital of Navarra (Pamplona, Spain) **(Table S1)**. Hyperprogressive disease was defined by radiological criteria and immunological profiles as described in Arasanz *et al* (*11*). The current prospective observational study was approved by the Ethics Committee of Clinical Investigations at the University Hospital of Navarre. Informed consent was obtained from all subjects and all experiments conformed to the principles set out in the WMA Declaration of Helsinki and the Department of Health and Human Services Belmont Report. Samples were collected through the Blood and Tissue Bank of Navarre, Health Department of Navarre, Spain. Eligible patients were 18 years of age or older who received immunotherapy targeting PD-1/PD- L1 as their current standard of care. Tumor PD-L1 expression was quantified in 103 of these patients. Age-matched healthy donors were recruited from whom written informed consents were obtained. The total number of donors was calculated *a priori* to ensure a power of 0.95 for F tests taking into consideration a large effect size (f=0.4). Power calculations were carried out with Gpower 3.1.9.7 (*65*).

Eight mL of peripheral blood samples were obtained prior and during immunotherapy before administration of each cycle. PBMCs were isolated as described (*2, 4, 66*) and myeloid cells analyzed by flow cytometry. The participation of each patient concluded when a radiological test confirmed response or progression, with the withdrawal of consent or after death of the patient. Tumor responses were evaluated according to RECIST 1.1 (*67*) and Immune-Related Response Criteria (*68*). Objective responses were confirmed by at least one sequential tumor assessment.

### Cell lines and ex vivo cell-based assays

3LL mouse adenocarcinoma cells were obtained from the American Type Culture Collection. Cells were grown in DMEM (Gibco) supplemented with 10% FBS and 1% penicillin/streptomycin following standard procedures. The murine NTCU-induced lung squamous cell carcinoma cell lines Lacun-3, UN-SCC679 and UN-SCC680 are described elsewhere (*50*). 5 human lung adenocarcinoma cell lines (Calu6, H23, A549, HCC44, H358) and 5 human lung squamous carcinoma cell lines (H520, H1703, H226, H2170, SW900) were obtained from the American Type Culture Collection and grown following standard procedures. When indicated, cell lines were engineered to constitutively overexpress soluble forms of GM-CSF, G-CSF, M-CSF and Fractalkine. Their coding sequences were synthesized (GeneArt Thermo Fisher) and cloned into pDUAL-Puromycin lentivectors (*69*) under the transcriptional control of the SSFV promoter. Lentivector production, titration, cell transduction and selection with puromycin were carried out as described elsewhere (*70*). Cytokine production was confirmed by ELISA. Cell growth and survival were monitored in real time using iCELLigence and xCELLigence real-time cell analysis (RTCA ACEA Biosciences) as described before (*70*). Scratch assays were carried out by seeding 1.5x10^5^ cells/well in 24 well plates with 100 nM of recombinant fractalkine (abcam) or 54 nM of the chemical inhibitor of the fractalkine receptor CX3CR1, AZD8797 (MedChemExpress). A scratch was made with a 1 mL pipette tip on a monolayer of growth-arrested cells. Bright field images were acquired at 0 and 48h timepoints with Cytation5 microscope (4X). ImageJ software was used to quantify the wound area.

### Flow cytometry

PBMC isolation, staining and flow cytometry were performed as described (*4, 70*). The following fluorochrome-conjugated anti-human antibodies were used at 1:50 dilutions unless otherwise stated: CD206 (15-2), CD124 (G077F6), LAG3 (11C3C65), CD38 (HB-7), CD69 (FN50), CD115 (9-4D2-1E4), PD-L1 (29E.2A3), C3AR (hC3Ar28, 1:200 diluted), CD64 (10.1, 1:100 diluted), CD32 (FUN-2, 1:500 diluted), CCR7 (G043H7), CD36 (5-271, 1:500 diluted), CD27 (M-T271), CD28 (CD28.2), CD8 (SK1) (Biolegend), CD163 (GHI/61.1), CD39 (REA739), TIM3 (F38-2E2), CD33 (AC104.3E3), VEGFR1 (REA569), CD16 (REA423), PD-1 (PD1.3.1.3), CD62L (145/15), CD10 (REA877), C5AR (S5/1), CD66B (REA306), CCR2 (REA624), CD56 (AF12-7H3), CD116 (REA211), CD4 (REA623), CD3 (REA613), CXCR1 (REA958), CXCR2 (REA2D8), CXCR4 (12G5), CX3CR1 (REA385) (Miltenyi), CD11b (M1/70, 1:300 diluted), CD14 (61D3, 1:20 diluted), CD86 (IT2.2), HLA-DR (L243, 1:25 diluted), CD54 (15.2), CD19 (SJ25C1), CD11c (3.9) (Tonbo), VISTA (B7H5DS8) (Invitrogen).

For immunophenotyping of circulating cell populations and tumor infiltrates in murine models, blood was retrieved from mice (50 µL) in EDTA-coated microtubes. Erythrocytes were lysed with BD FACS lysing solution for 1 minute and the resulting cell suspensions were stained and analyzed by flow cytometry. Tumors were harvested and mechanically disaggregated. Erythrocytes were lysed and the single-cell suspensions were stained and analyzed by flow cytometry. The following fluorochrome-conjugated anti-mouse antibodies were used: Ly6C (REA796), F4/80 (REA126) (Miltenyi), Ly6G (1A8), CD115 (AFS98), MHC-II (M5/114.15.2), CX3CR1 (SA011F11), CCR2 (SA203G11), NK1.1 (PK136), CD4 (GK1.5), CD8 (53-6.7) (Biolegend), CD45 (30-F11, 1:250 diluted), CD11c (HL3) (BD Pharmigen), CD11b (M1/70), CD3 (145-2C11) (Tonbo), CD45 (104, 1:250 diluted), CD19 (eBio1D3) (eBioscience), CD25 (7D4) (Southern biotech).

All samples were acquired in a FACS Canto II flow cytometer (Becton Dickinson). Flow cytometry data were exported as FCS3.0 files and analyzed using FlowJo or SPADE software. Gating strategies are shown in **Fig.S10**.

The clustering algorithm SPADE (spanning tree progression analysis of density-normalized events, Stanford University) was used to integrate and analyze multiple flow cytometry panels using internal FSC-SSC patterns and common CD11b and CD14 as overlapping markers to reconstruct SPADE trees (*39*). Diversity indexes were defined as the total number of terminal branches within each hierarchical cluster tree.

### Cytokine quantification

Human plasma samples were obtained from 8 ml-EDTA blood tubes from each patient. Murine blood samples were collected by facial vein puncture in EDTA coated microtubes and plasma obtained by standard procedures. Quantification of plasma soluble cytokines, chemokines and soluble immune checkpoints concentrations in plasma samples was carried out by luminex xMAP technology following manufacturer’s instructions. A Human Cytokine/Chemokine magnetic bead panel (HCYTOMAG-60K, Millipore) was used to measure the concentrations of CD40L, CCL11, IFNα, IFNγ, IL2, IL4, IL6, IL8, IL12, IL17, TNFα, VEGFA, FGF2, FKN, GCSF, GMCSF, IL1β, IL1Ra, IL3, IL5, IL7, IL9, IL10, IL15, CXCL10, MCP1, CCL7, CCL3 and CCL4. A human Immuno-oncology checkpoint protein magnetic bead panel (HCKPMAG- 11K, Millipore) was used to quantify concentrations of soluble BTLA, CD27, CD28, TIM3, HVEM, CD40, GITR, LAG3, TLR2, GITRL, PD1, CTLA4, CD80, CD86, PDL1 and ICOS. A

Human Immuno-oncology checkpoint protein magnetic bead panel 2 (HCKP2-11K, Millipore) was used to quantify concentrations of soluble arginase-1, ICOSL, CD276, CD73, VTCN1, APRIL, VISTA, B7-H6, granzyme B, E-cadherin, galectin-1, galectin-3, granulysin, IDO1, MIC-A, MIC-B, BAFF, OX40, CD155 and perforin. Final detection and data analyses were performed on a MAGPIX (EMD Millipore) with xPONENT software.

Quantification of plasma soluble FKN, IFNγ, IL10, IL12p70 and IL4 in murine plasma samples was carried out by ELISA following the manufacturer’s instructions (R&D DuoSet ELISA kits; DY472 for FKN; DY485-05 for IFNγ; DY417-05 for IL10; DY419-05 for IL12p70 and DY404-05 for IL4). Quantification of secreted M-CSF, G-CSF, FKN and GM-CSF from cell cultures were carried out by ELISA following standard procedures (R&D DuoSet ELISA kits; DY415-05 for GM-CSF; DY414-05 for GCSF; DY416-05 for MCSF; MCX310 for FKN). Secreted FKN concentration was quantified in supernatants of human cancer cell lines by ELISA (DY365 R&D DuoSet ELISA kit).

### Mouse lung cancer models and therapies

Approval for animal studies was obtained from the Animal Ethics Committee of the University of Navarra (Pamplona, Navarra, Spain. Reference 077-19 and 049-18) and from the Government of Navarra. The indicated 3LL cell lines (1.5x10^6^ cells/mouse; n=6 mice/group) were subcutaneously injected in the flanks of 10-week old C57BL/6 female mice (Envigo). 3LL engraftments were allowed to grow for 7 days, then mice were intraperitoneally treated with 100 µg of anti-PD-1 (RPMI-14, BioXCell) or saline buffer as control vehicle at days 7, 11 and 15 after tumor inoculation. Tumor size was measured three times per week with a digital caliper until humane endpoint was reached (tumor large diameter superior than 14 mm). Tumor volumes (V) were calculated as V (mm^3^) = [(short diameter)^2^ x (long diameter)]/2. Analyses of circulating immune populations were carried out at day 15 after tumor engraftment. For immunophenotyping of tumor infiltrates, tumors were harvested and mechanically disaggregated 14 days after engraftment (1.5x10^6^ cells/mouse; n=8 mice/group). To assess abscopal effects of systemic FKN, 3LL or 3LL-FKN cells were subcutaneously injected (1.5x10^6^ cells/mouse; n=6 mice/group) on the left flank of mice. 7 days after tumor engraftment, 3LL cells were injected on the right flank. 3LL engraftments were allowed to grow for 7 days, then mice were intraperitoneally treated with 100 µg of anti-PD-1 (RPMI-14, BioXCell) or saline buffer as control vehicle at days 7, 11 and 15 after tumor inoculation. *In vivo* NK, CD4 and CD8 T cell depletions were carried out by intraperitoneal administration of 100 µg of anti-mouse CD8a (clone 2.43; BioXCell), CD4 (clone GK1.5; BioXCell) or NK1.1 (clone PK136; BioXCell) antibodies, respectively, at days 6, 10, 14 and 18 after 3LL tumor inoculation.

In some experiments, intraperitoneal administration of 100 µg/Kg of recombinant FKN was performed (abcam) 6 days after tumor inoculation (1.5x10^6^ cells/mouse; n=6 mice/group) in 10- week old C57BL/6 male mice (Envigo). When indicated, intraperitoneal injection the CX3CR1 chemical inhibitor AZD8797 (MedChemExpress) was carried out with 0.8 mg/Kg at days 6, 8, 11, 13 and 15 after tumor engraftment.

To study in vivo tumor growth of the lung squamous cell line UN-SCC680, 1.5x10^6^ cells/mouse were subcutaneously administered (n=6 mice/group) to 10-week old female A/JOlaHsd (A/J) mice (Envigo). Tumor engraftments were allowed to grow for 7 days, then mice were intraperitoneally treated with 100 µg of anti-PD-1 (RPMI-14, BioXCell) or saline buffer as control vehicle at days 6, 11 and 15 after tumor inoculation.

### Transcriptomic data analyses

To assess the association of FKN mRNA expression with infiltration of different immune populations in human lung adenocarcinomas, we used Tumor Immune Estimation Resource or TIMER2.0 (http://timer.cistrome.org/) (*44*). This resource contains transcriptional data from samples included in the TCGA database, including the abundance of immune cells across multiple tumor types. It generates a graphical representation of user-selected gene expression and Spearman correlations with immune populations of interest as identified by CIBERSORT, quanTIseq, xCell and TIDE algorithms. We restricted our analysis to lung adenocarcinomas and performed systematic correlation studies with CD8 lymphocytes, CD4 lymphocytes, regulatory T lymphocytes, B cells, NK cells, neutrophils, monocytes, macrophages, dendritic cells and MDSCs. A partial purity-adjusted Spearman’s correlation was used.

To evaluate the prognostic value of FKN in this database, Cox proportional hazards model was performed, with graphical representations of Kaplan-Meier overall survival with Gene Expression Profiling Interactive Analysis, or GEPIA2 (http://gepia2.cancer-pku.cn) (*45*). Groups were distributed according to the median. Correlation between FKN and PD-L1 expression was performed by Spearman correlation analyses within this analysis platform.

### Statistical analysis and study design

No data was considered an outlayer, or removed from the analyses. For mouse experiments, sample sizes were calculated to achieve a minimum power of 0.8 for F-based tests taking into consideration a large effect size (f=04). Power calculations were carried out with Gpower 3.1.9.7. All variables were tested for normality using the Kolgomorov-Smirnov test. Homogeneity was assessed with Spearman’s coefficient of variations. Samples were considered homogeneous with CV<25%. Homogeneity of variances was tested by the F-Fisher test. Homogeneous groups with comparable variances and fulfilling normality were compared using parametric tests. For multicomparisons, one-way ANOVAs were used followed by pairwise comparisons with *a posteriori* Tukey’s tests. Variables fulfilling these criteria were (1) diversity indexes as calculated below; (2) cell growth rates and cell index data from ACEA Real-Time Cell monitoring (RTCA) data; (3) Tumor volumes as measured at the indicated time points; (4) FKN plasma concentrations in tumor-bearing mice, and (5) secreted FKN concentrations from cultures of cancer cell lines.

The rest of variables were tested with non-parametric tests. For multicomparisons, one-way Kruskal-Wallis tests were used followed by *a posteriori* pair-wise comparisons with Dunn’s test. Variables that were normally distributed but did not fulfil homogeneity of variances were also tested with non-parametric tests. These included (1) percentages of monocytes and neutrophils quantified by high-dimensional flow cytometry; (2) human serum cytokine concentrations, and (3) percentages of circulating and tumor-infiltrating immune cell types quantified by flow cytometry, in mouse tumor models.

DI in circulating CD11b+ cells were calculated as the number of clusters with terminal phenotypes from SPADE 3 hierarchical cluster analyses using high-dimensional flow cytometry data. Correlations between DI and other variables (which included percentages of monocytes, neutrophils and serum concentrations of relevant factors) were evaluated by Spearman’s tests. The predictive capacities of the selected indicated variables were assessed by ROC analyses as described before (*4*). PFS and OS in human patients and in mouse tumor models were compared by Log-rank tests.

## List of Supplementary Materials

**Fig. S1.**
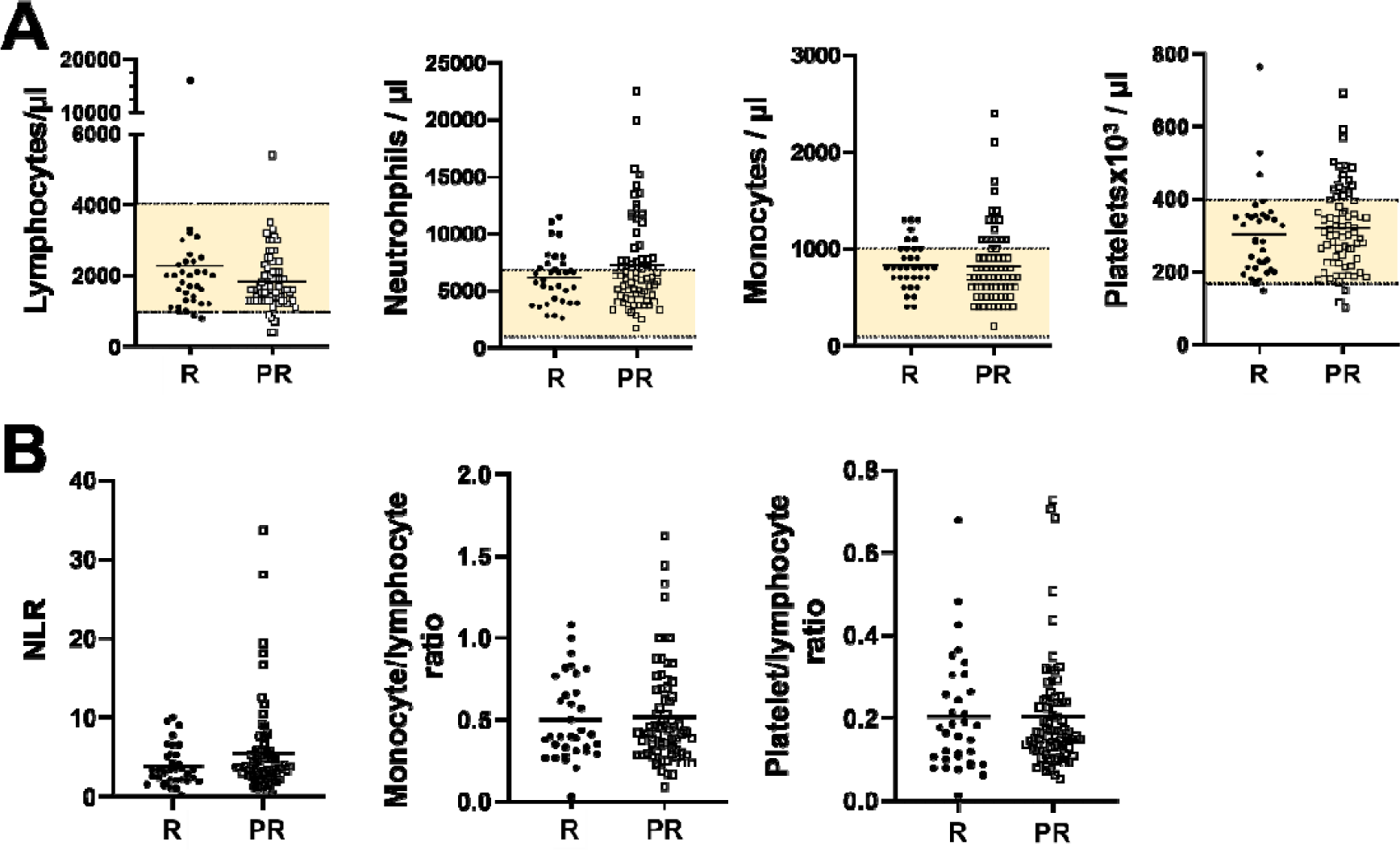
Quantification of major immune cell populations by standard analytical clinical techniques and correlates with response to immunotherapy. (**A**) Dot plot of the indicated immune cell counts in the indicated patient groups before immunotherapies. Yellow shaded areas indicate the normal range of analyzed blood populations. (**B**) NLR, monocyte/lymphocyte ratio and platelet/lymphocyte ratio in the indicated patient groups before immunotherapies. R, responder patients; PR, non-responder patients. No statistical differences were observed between groups.

**Fig S2.**
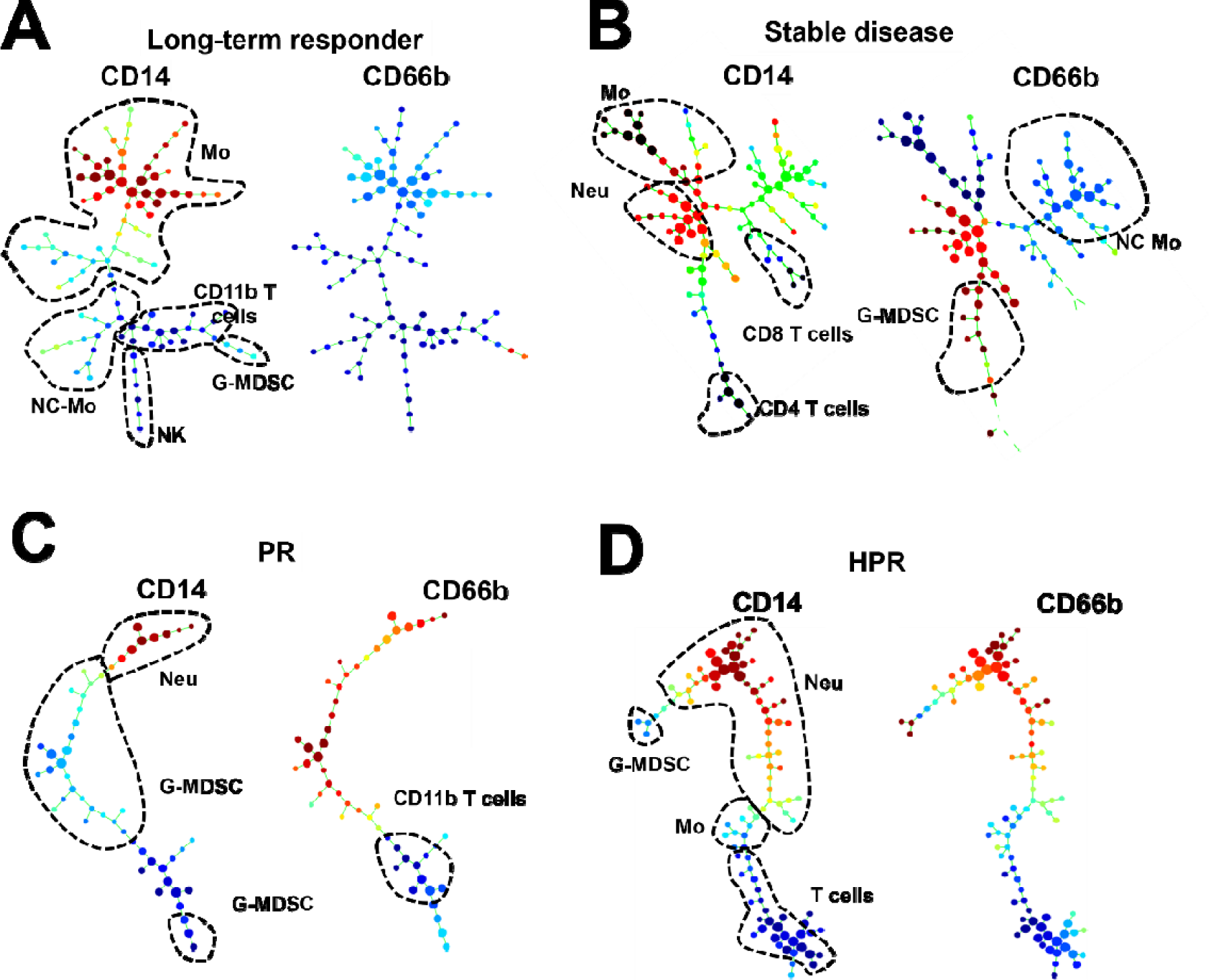
Hierarchical phenotype clustering from high dimensional flow cytometry data of baseline multiple cell lineage and activation markers within immune cell subsets in PBMC from NSCLC patients undergoing anti-PD-1/PD-L1 immunotherapy. Representative SPADE3 cluster profiles of myeloid cells integrating 43 markers are shown for (**A**) a long term responder, **(B)** a short term responder (stable disease), **(C)** a progressor (PR) and **(D)** a hyperprogressor (HPR). Distributions of CD14 (left dendrogram) and CD66b expression (right dendrogram) are shown. Main cell subsets are encircled and identified as Mo, monocytes; Neu, neutrophils; G- MDSC, granulocytic myeloid-derived suppressor cells; NC-Mo, non-classical monocytes; NK, natural killer cells. The relative expression of the selected marker as indicated above the graphs color-coded, from dark red (maximum expression) to dark blue (minimum expression).

**Figure S3.**
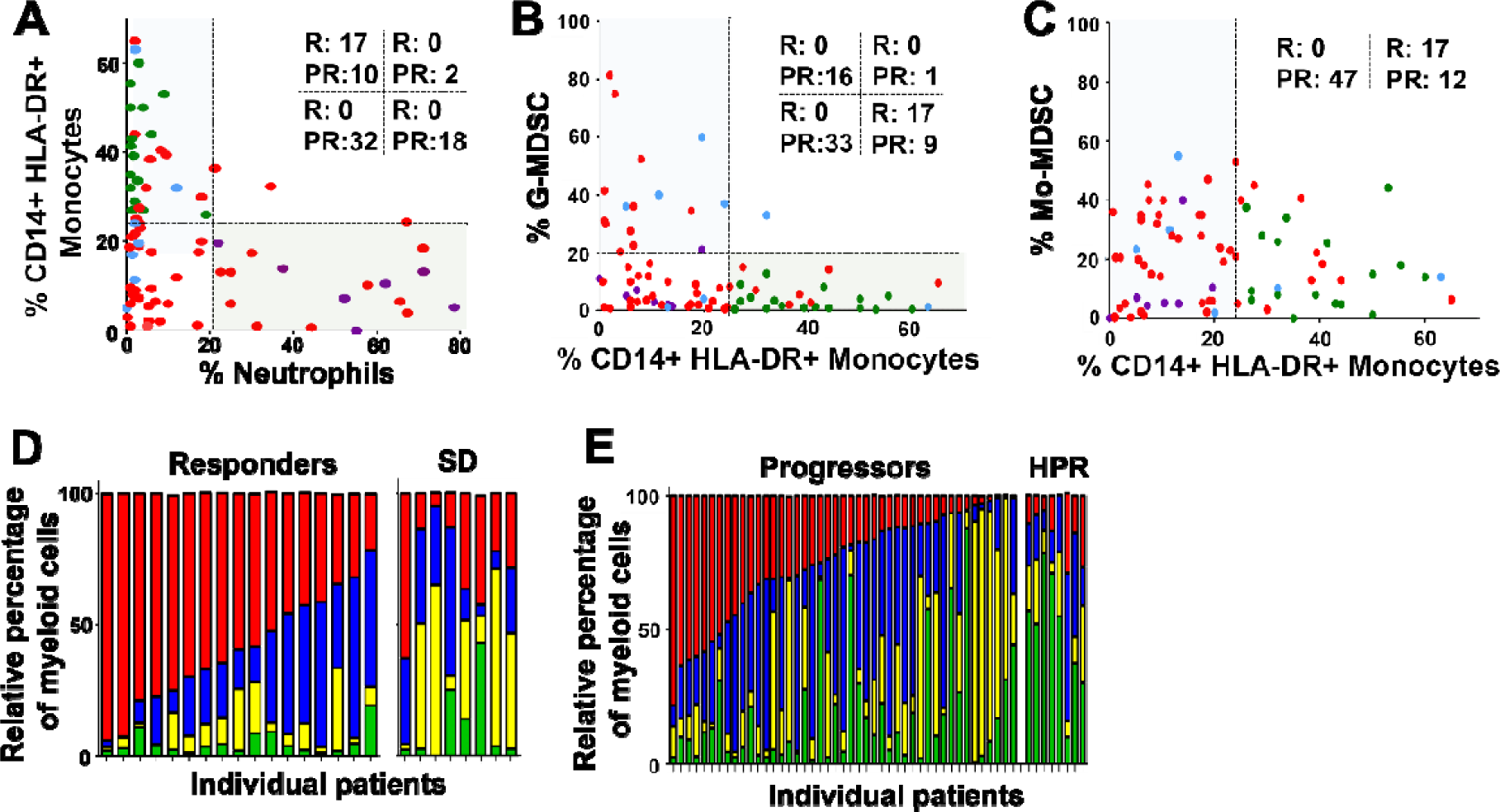
Baseline monocyte/neutrophil profiles in peripheral blood from NSCLC patients undergoing PD-L1/PD-1 blockade. (**A**) The baseline frequency of monocytes (CD14^+^ HLA- DR^+^) vs neutrophils (CD14^+^ CD66b^+^) was analyzed within CD11b^+^ cells in patients classified as responders (R, green), progressors (PR, red), stable disease (blue) and hyperprogressors (purple). The number of responders and progressors is indicated in each quadrant. **(B)** As in (A) but plotting the percentage of G-MDSCs (CD14^-^ CD66b^+^) vs monocytes. **(C)** As in (A) but plotting the percentage of Mo-MDSC (CD14^+^ HLA-DR^-^) vs monocytes. **(D)** The relative percentages of the main myeloid populations restricted to this compartment is plotted for each patient under study as indicated as color codes, classified according to objective responders and stable disease (SD). **(e)** As in (D) but in progressors and hyperprogressors (HPR).

**Fig. S4.**
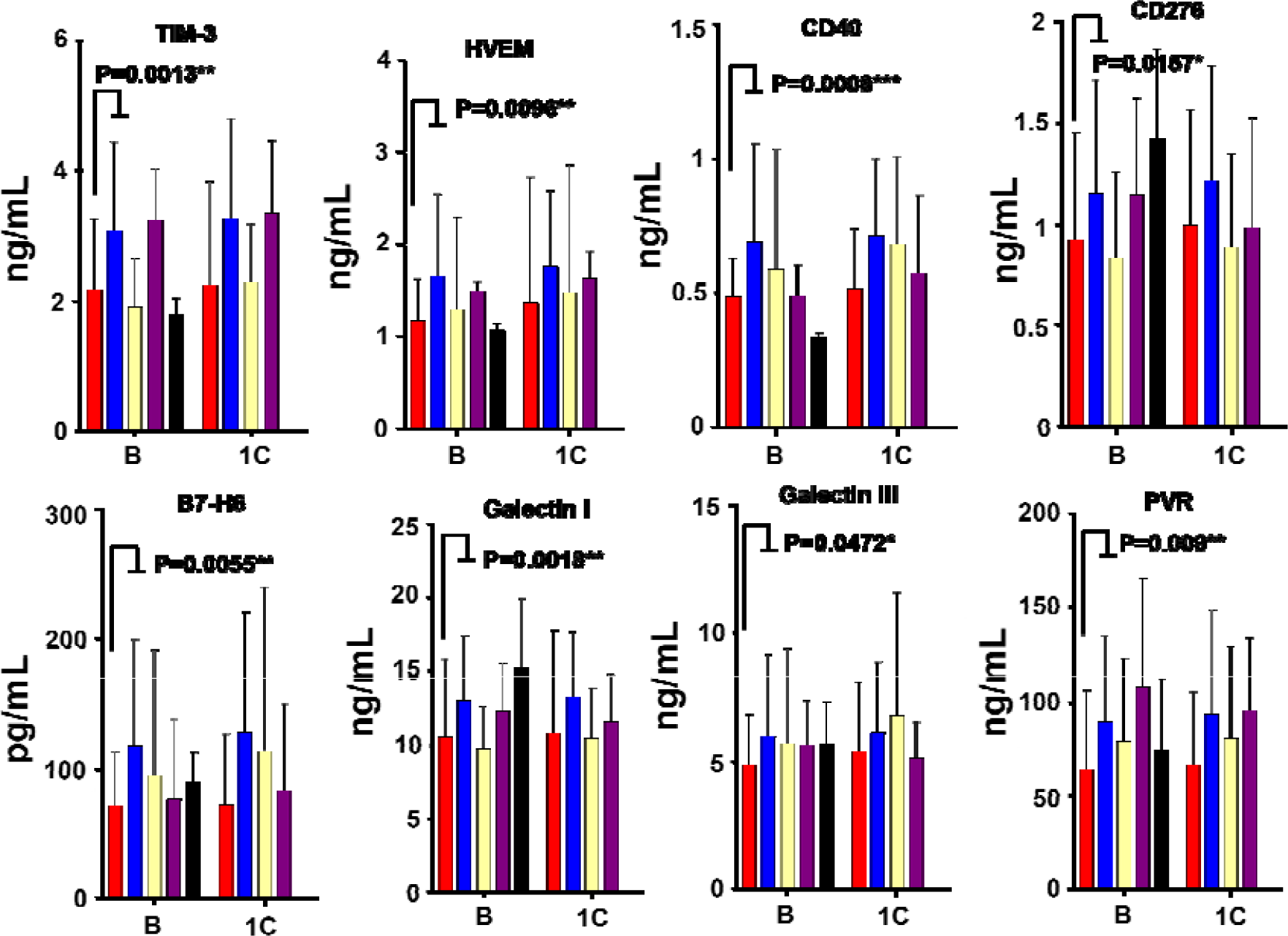
Quantification of plasma soluble factors with broad immunomodulatory properties. The bar graphs show plasma concentrations of the indicated proteins in responders (R, n=27, 28), progressors (PR, n=59, 63), stable disease (SD, n=11, 13), hyperprogressors (HPR, n=3) and age-matched healthy donors (H, n=4, 6) before (baseline, B) and after the first cycle of immunotherapy (1C). Error bars are shown (SD), and relevant statistical comparisons as shown within the graphs were carried out by the test of Wilcoxon followed (when relevant) by pair-wise comparisons with the Mann-Whitney U test. *, **, ***, indicate significant (p<0.05), very significant (p<0.01) and highly significant (p<0.001) differences.

**Fig. S5.**
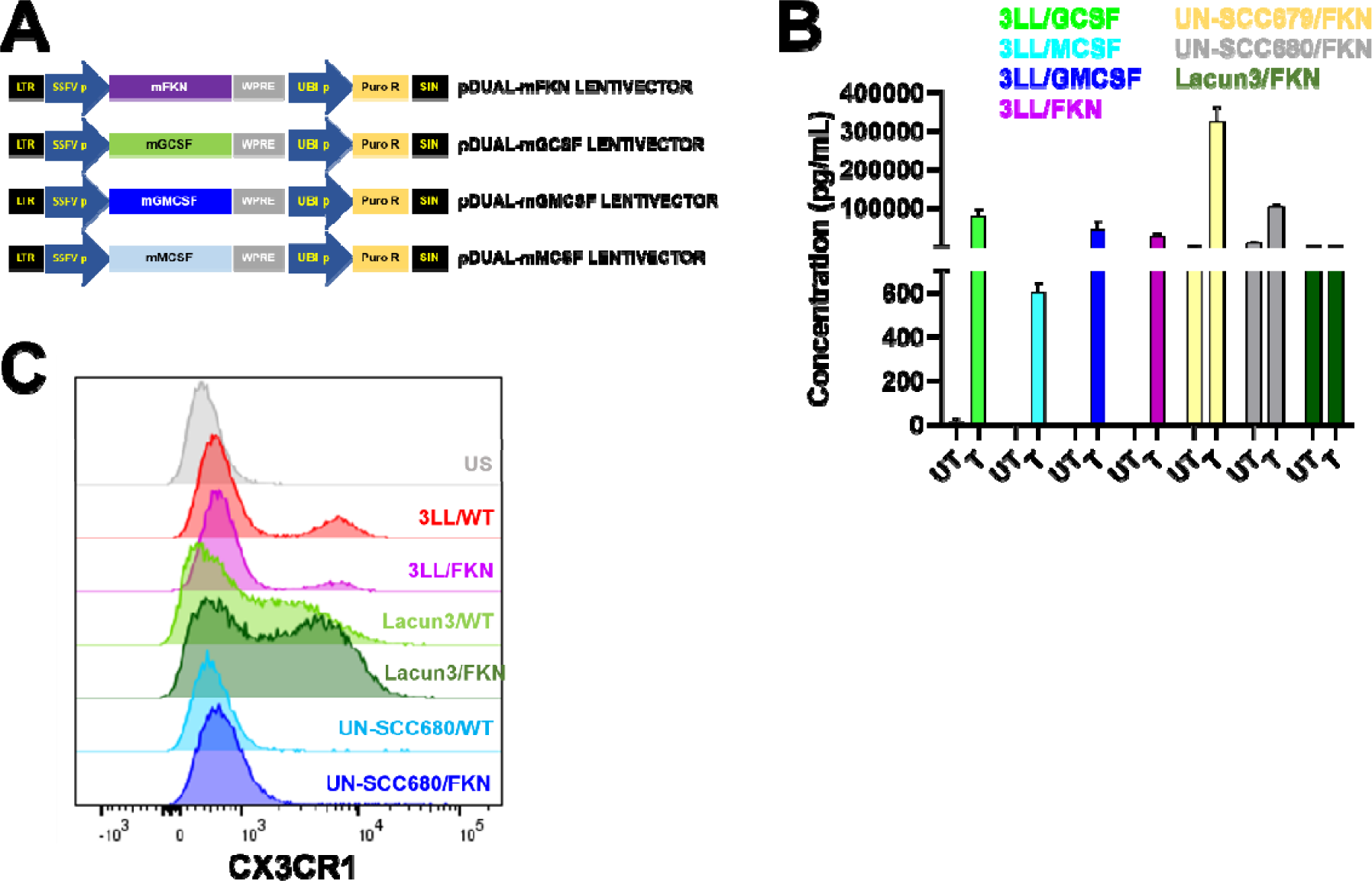
Characterization of the engineered lung cancer cell lines overexpressing selected myeloid-regulating cytokines. **(A)** Structure of the lentivectors used to engineer lung cancer cell lines, with the relevant murine cytokine genes as shown in each lentivector. SIN, self-inactivating deleted LTR; LTR, long-terminal repeat; SFFVp, spleen focus-forming virus promoter; WPRE, woodchuck post-transcriptional regulatory element; UBIp, human ubiquitin promoter; Puro R, puromycin resistance gene. **(B)** ELISA quantification of cytokine secretion by the indicated engineered LUAD and LUSC lung cancer cell lines (T) to overexpress the selected cytokines as shown. Endogenous secretion of each cytokine was also tested by ELISA quantification in supernatants from cultures of parental unmodified controls (UT). **(C)** Flow cytometry histogram plots of FKN receptor CX3CR1 surface expression in the indicated LUAD and LUSC lung cancer cell lines, either parental (WT) or modified to overexpress FKN.

**Fig. S6.**
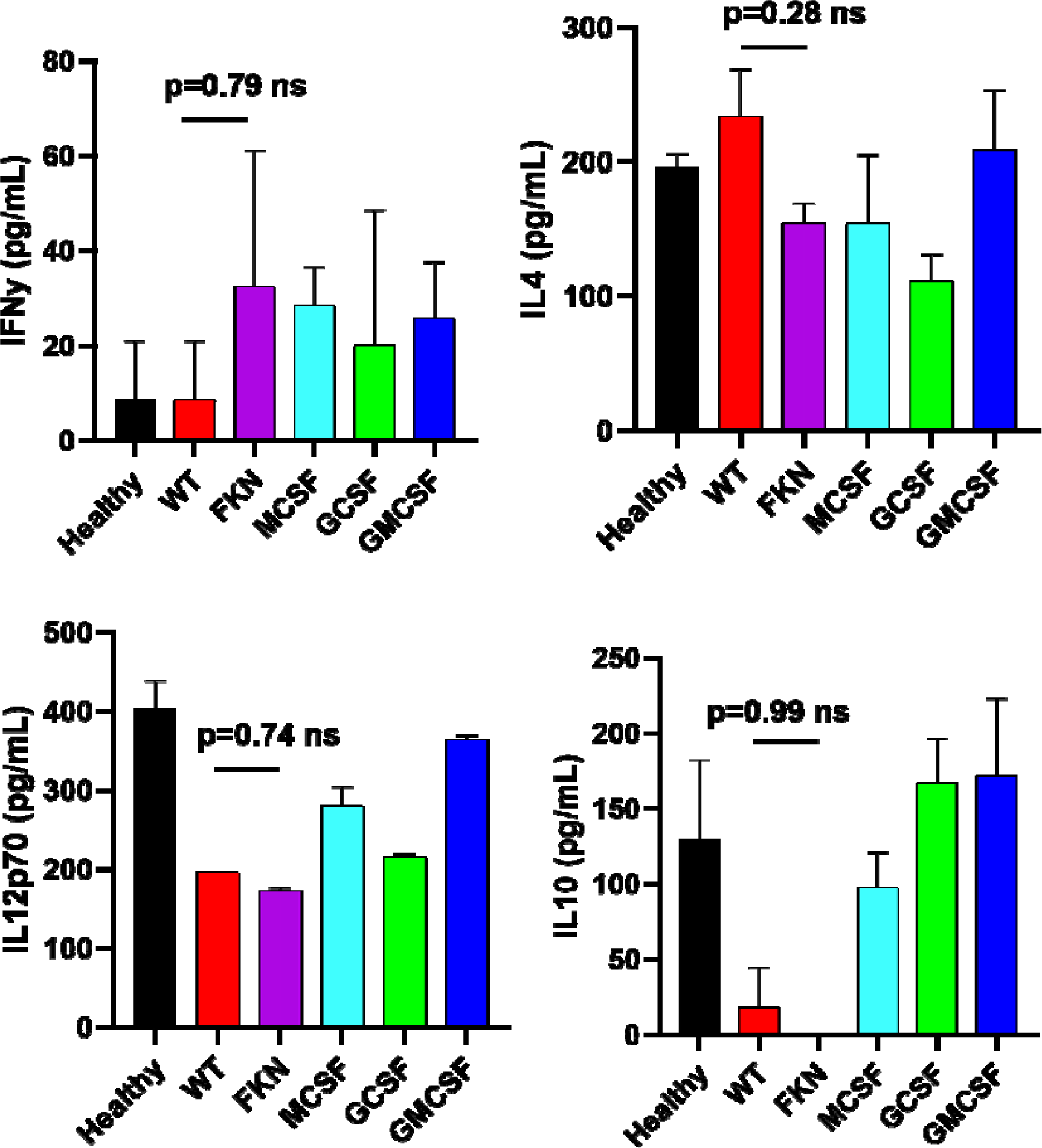
Quantification of systemic cytokines in mice transplanted with cytokine-expressing 3LL tumors treated with anti-PD1 blockade therapy. Plasma concentrations of pro- and anti- inflammatory cytokines at day 15 after tumor inoculation. Data are expressed as mean ± SD from a pool of 6 mice/group. Relevant statistical comparisons were performed by Wilcoxon’s test followed by pair-wise comparisons of relevance by the Mann-Whitney U test. ns, non-significant differences.

**Fig. S7.**
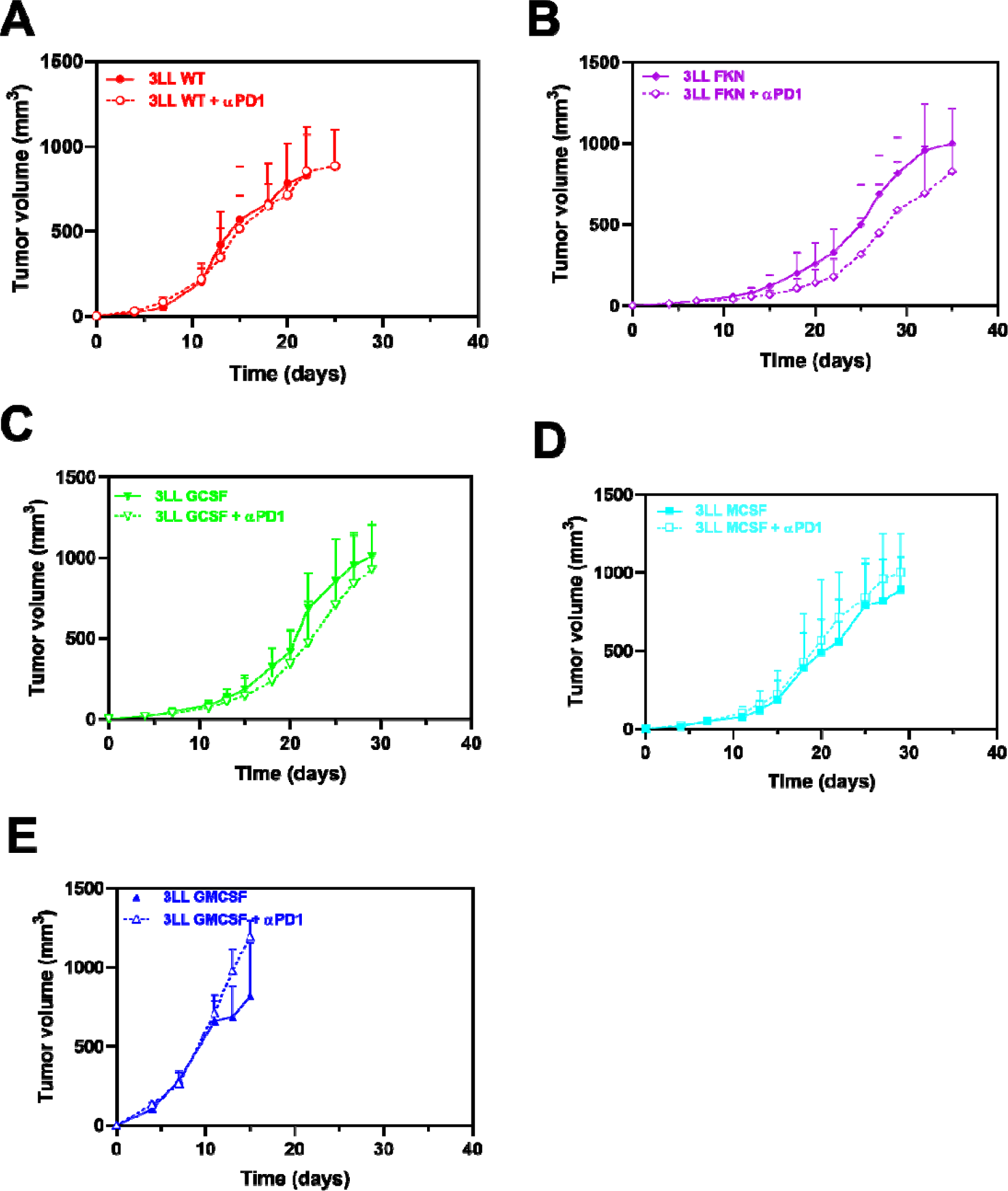
Anti-tumor synergy of secreted fractalkine with anti-PD-1 blockade. (**A**) Tumor growth of the parental 3LL cell line (WT) transplanted into groups of mice (n=6) with or without PD-1 blockade, as indicated. Mice were treated with either anti-PD-1 antibody (αPD1) or control vehicle (saline buffer) at days 7, 11 and 15 after tumor inoculation. Data are shown as means ± SD. **(B)** As in (A) but with FKN-expressing 3LL cells. **(C)** As in (A) but with GCSF- expressing 3LL cells; **(D)** As in (A) but with MCSF-expressing 3LL cells; **(e)** As in (A) but with GMCSF-expressing 3LL cells.

**Fig. S8.**
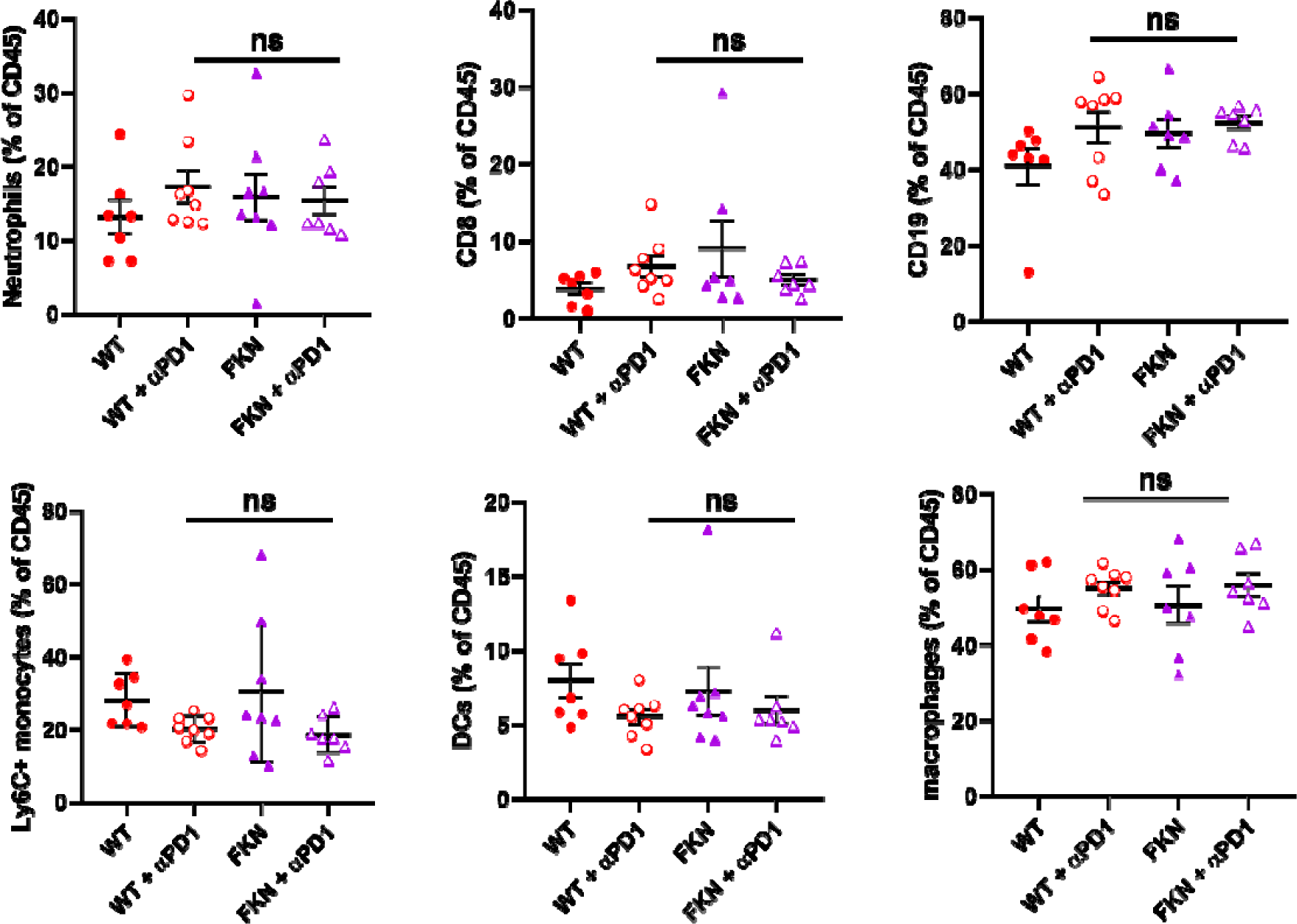
Tumor infiltrating immune cells in mice transplanted with 3LL cells expressing FKN and its combination with PD-1 blockade. The graphs represent percentages of the indicated infiltrating immune cell types as quantified by flow cytometry, in tumors excised from mice inoculated with the indicated cell lines (parental cell line, WT; 3LL cells expressing FKN, FKN) with or without PD-1 blockade. Neutrophils, CD8, B cells (CD19), Ly6C^+^ monocytes, DCs (CD11c) and macrophages (F4/80) were quantified at day 14 after tumor inoculation. Data are shown as the mean of the percentage within total leukocytes (CD45^+^) ± SD (n=8 mice). Relevant statistical comparisons are shown within the graphs, evaluated by ANOVA and Tukey’s pairwise comparisons. ns, non-significant differences.

**Fig. S9.**
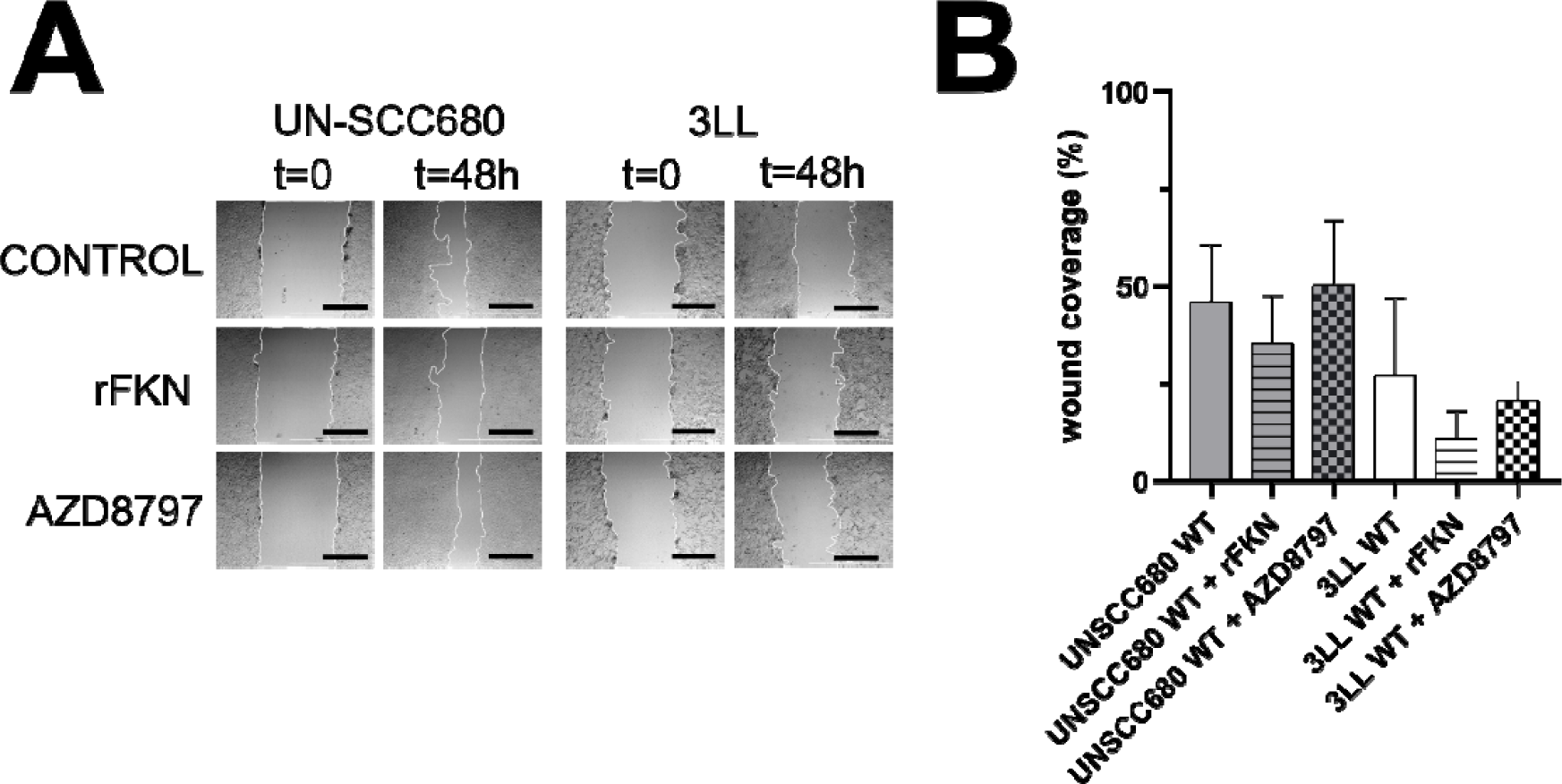
In vitro invasion potential of lung cancer cells of different histologies. (**A**) Assessment of the invasion potential of the murine LUSC model cell line UN-SCC680 and the LUAD cell line 3LL by the scratch assay, treated with recombinant fractalkine (rFKN) or the allosteric non- competitive antagonist of the FKN receptor CX3CR1 (AZD8797), compared to untreated controls. Phase contrast microscopy pictures are shown, taken at the time intervals indicated on top. Scale bars, 1 mm. **(B)** Bar graph presenting the percentage of the area covered by migrating cells (wound coverage), expressed as mean ± SD (n=3 independent replicates).

**Fig. S10.**
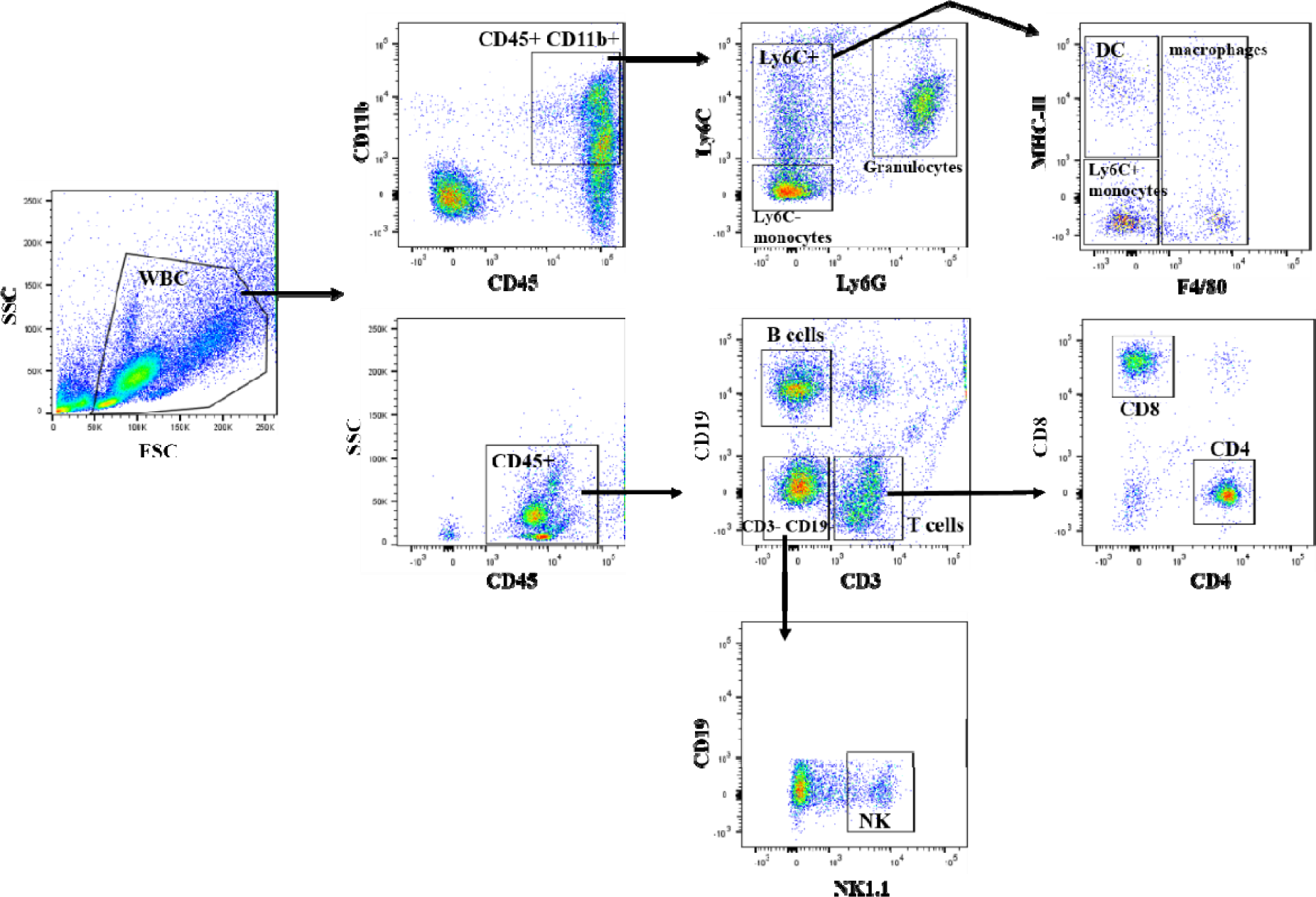
Gating strategy followed to analyze FACS data. Murine samples were processed and erythrocytes were lysed to isolate white blood cells (WBC). Samples were stained with fluorochrome-conjugated antibodies and acquired on an FACS Canto II flow cytometer. Data were exported as FCS3.0 files and analyzed using FlowJo software.

**Supplementary table 1.** Patient cohort characteristics.

| Variable | All patients (n=112) |
| --- | --- |
| **Sex** |  |
| Female | 28 (25.0 %) |
| Male | 84 (75.0 %) |
| **Age** |  |
| <60 | 31 (27.7 %) |
| ≥60 | 81 (72.3 %) |
| **Histology** |  |
| Squamous | 28 (25.0 %) |
| Non- Squamous | 84 (75.0 %) |
| **Immunotherapy treatment** |  |
| Pembrolizumab (αPD-1) | 32 (28.6 %) |
| Nivolumab (αPD-1) | 34 (30.3 %) |
| Atezolizumab (αPD-L1) | 33 (29.5 %) |
| Pembrolizumab + platinum-based chemotherapy | 13 (11.6 %) |
| **Tumor PD-L1 status** |  |
| 0% | 38 (34.0 %) |
| 1-4% | 12 (10.7 %) |
| 5-49% | 20 (17.8 %) |
| ≥ 50% | 33 (29.5 %) |
| Undetermined | 9 (8.0 %) |
| **Mutation status** |  |
| Not evaluated | 108 (96.4 %) |
| EGFR | 1 (0.9 %) |
| ROS1 | 1 (0.9 %) |
| KRAS | 1 (0.9 %) |
| METamp | 1 (0.9 %) |
| **Smoking status** |  |
| Smoker | 106 (94.6 %) |
| Non-smoker | 6 (5.4 %) |
| **Treatment line** |  |
| 1st | 38 (33.9 %) |
| 2nd | 61 (54.5 %) |
| 3th | 10 (8.9 %) |
| 4th or higher | 3 (2.7 %) |
| **Previous systemic therapies (previous 3 months)** |  |
| Platinum-based therapy | 28 (25.1 %) |
| Non-platinum based therapy | 36 (32.1 %) |
| No | 48 (42.8 %) |
| **Liver metastases** |  |
| No | 83 (74.1 %) |
| Yes | 29 (25.9 %) |
| **Number of metastatic sites** |  |
| ≤2 | 33 (29.5 %) |
| ≥3 | 79 (70.5 %) |
| **Responses** |  |
| Partial response | 29 (25.9 %) |
| Progression disease | 65 (58.0 %) |
| Hyperprogression | 5 (4.5 %) |
| Stable disease | 13 (11.6 %) |

**Supplementary table 2.**
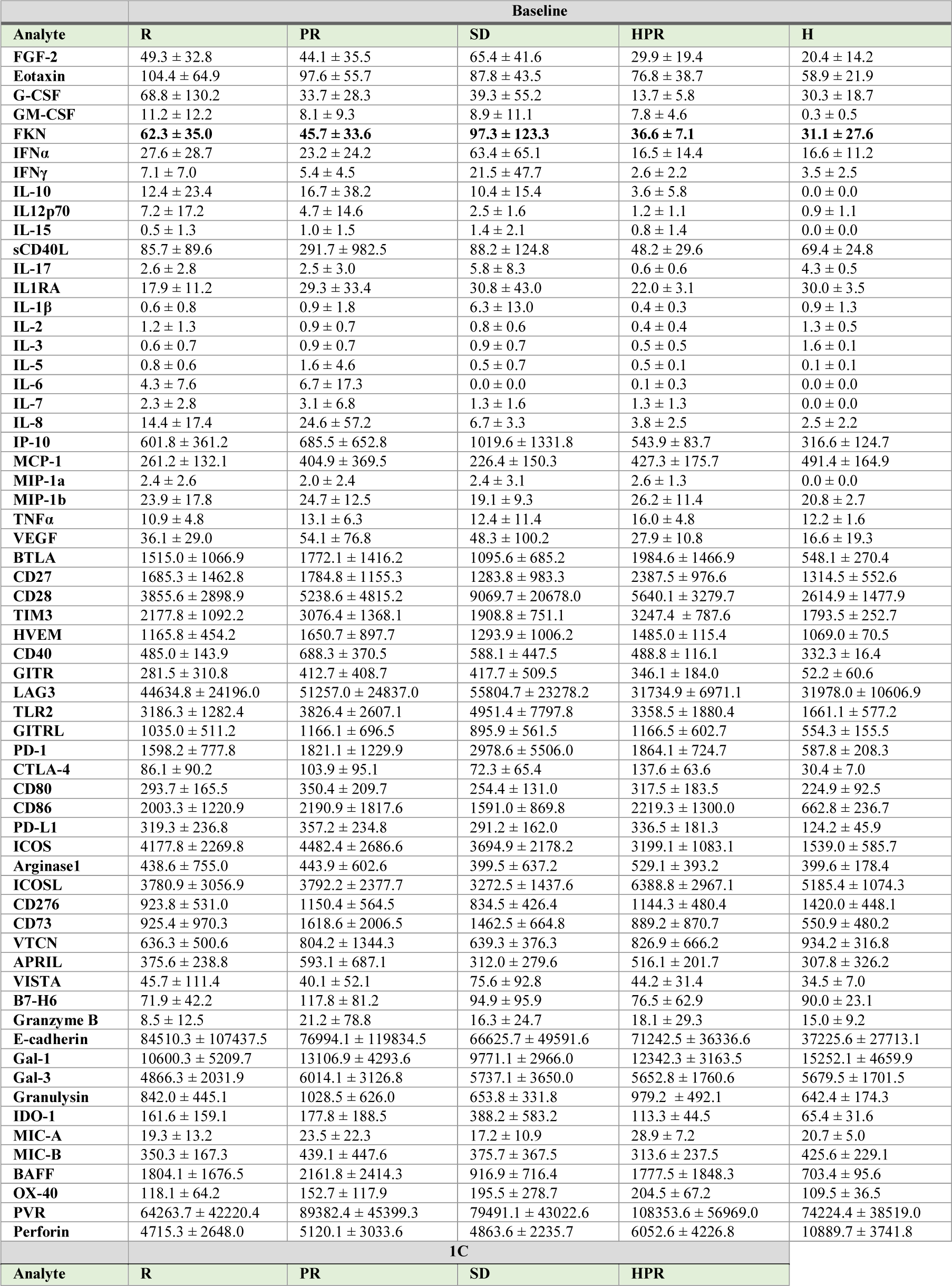
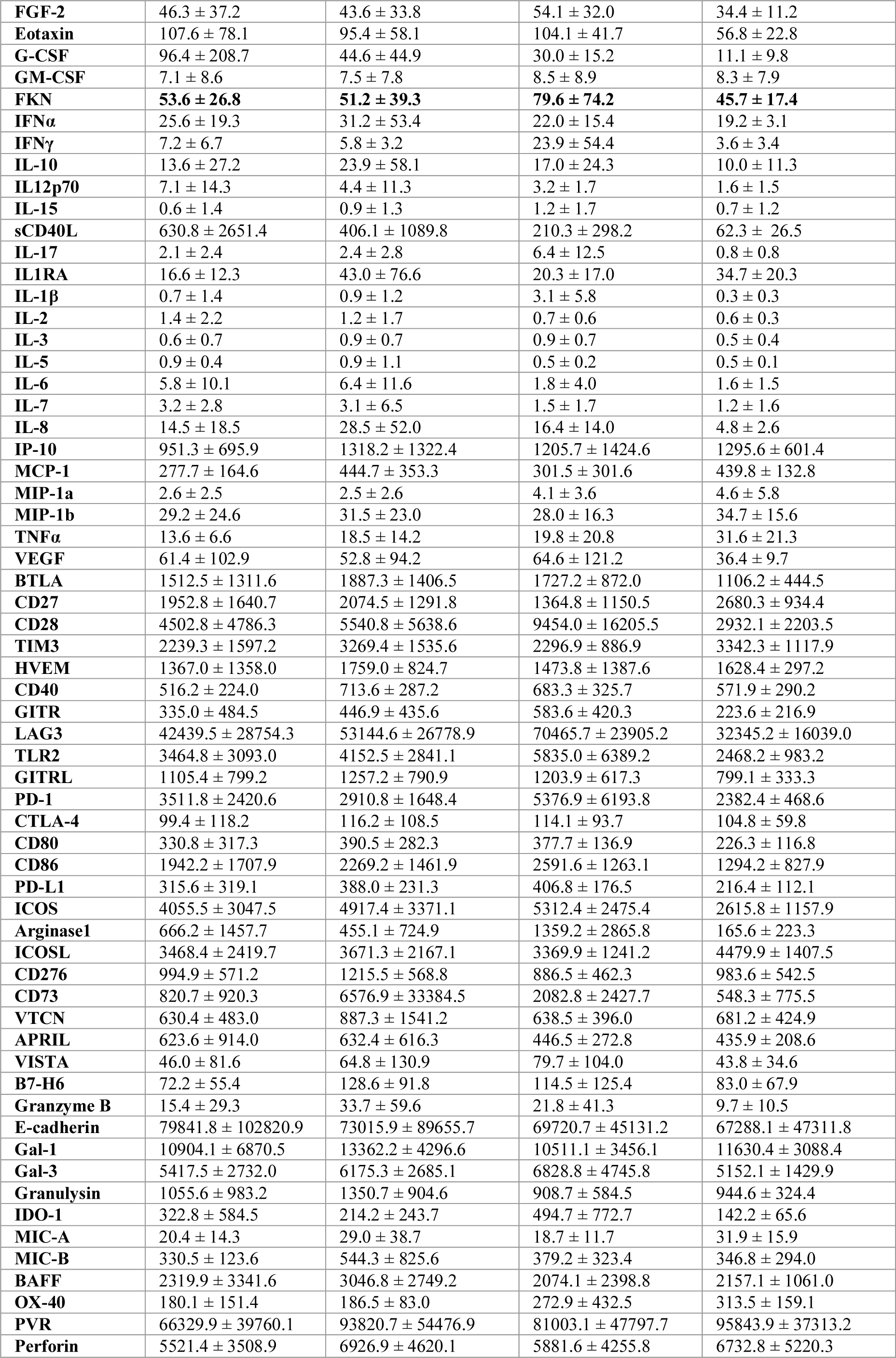
Concentration of plasma soluble-proteins quantified by luminex prior to immunotherapy (Baseline) and after the first cycle of treatment (1C). Data are expressed as mean (pg/mL) ± SD. R: responders; PR: non-reponders; SD: stable disease; HPR: hyperprogressors; H: age-matched healthy donors.

|  | Baseline |  |  |  |  |
| --- | --- | --- | --- | --- | --- |
| Analyte | R | PR | SD | HPR | H |
| FGF-2 | 49.3 ± 32.8 | 44.1 ± 35.5 | 65.4 ± 41.6 | 29.9 ± 19.4 | 20.4 ± 14.2 |
| Eotaxin | 104.4 ± 64.9 | 97.6 ± 55.7 | 87.8 ± 43.5 | 76.8 ± 38.7 | 58.9 ± 21.9 |
| G-CSF | 68.8 ± 130.2 | 33.7 ± 28.3 | 39.3 ± 55.2 | 13.7 ± 5.8 | 30.3 ± 18.7 |
| GM-CSF | 11.2 ± 12.2 | 8.1 ± 9.3 | 8.9 ± 11.1 | 7.8 ± 4.6 | 0.3 ± 0.5 |
| FKN | **62.3 ± 35.0** | **45.7 ± 33.6** | **97.3 ± 123.3** | **36.6 ± 7.1** | **31.1 ± 27.6** |
| IFNα | 27.6 ± 28.7 | 23.2 ± 24.2 | 63.4 ± 65.1 | 16.5 ± 14.4 | 16.6 ± 11.2 |
| IFNγ | 7.1 ± 7.0 | 5.4 ± 4.5 | 21.5 ± 47.7 | 2.6 ± 2.2 | 3.5 ± 2.5 |
| IL-10 | 12.4 ± 23.4 | 16.7 ± 38.2 | 10.4 ± 15.4 | 3.6 ± 5.8 | 0.0 ± 0.0 |
| IL12p70 | 7.2 ± 17.2 | 4.7 ± 14.6 | 2.5 ± 1.6 | 1.2 ± 1.1 | 0.9 ± 1.1 |
| IL-15 | 0.5 ± 1.3 | 1.0 ± 1.5 | 1.4 ± 2.1 | 0.8 ± 1.4 | 0.0 ± 0.0 |
| sCD40L | 85.7 ± 89.6 | 291.7 ± 982.5 | 88.2 ± 124.8 | 48.2 ± 29.6 | 69.4 ± 24.8 |
| IL-17 | 2.6 ± 2.8 | 2.5 ± 3.0 | 5.8 ± 8.3 | 0.6 ± 0.6 | 4.3 ± 0.5 |
| IL1RA | 17.9 ± 11.2 | 29.3 ± 33.4 | 30.8 ± 43.0 | 22.0 ± 3.1 | 30.0 ± 3.5 |
| IL-1β | 0.6 ± 0.8 | 0.9 ± 1.8 | 6.3 ± 13.0 | 0.4 ± 0.3 | 0.9 ± 1.3 |
| IL-2 | 1.2 ± 1.3 | 0.9 ± 0.7 | 0.8 ± 0.6 | 0.4 ± 0.4 | 1.3 ± 0.5 |
| IL-3 | 0.6 ± 0.7 | 0.9 ± 0.7 | 0.9 ± 0.7 | 0.5 ± 0.5 | 1.6 ± 0.1 |
| IL-5 | 0.8 ± 0.6 | 1.6 ± 4.6 | 0.5 ± 0.7 | 0.5 ± 0.1 | 0.1 ± 0.1 |
| IL-6 | 4.3 ± 7.6 | 6.7 ± 17.3 | 0.0 ± 0.0 | 0.1 ± 0.3 | 0.0 ± 0.0 |
| IL-7 | 2.3 ± 2.8 | 3.1 ± 6.8 | 1.3 ± 1.6 | 1.3 ± 1.3 | 0.0 ± 0.0 |
| IL-8 | 14.4 ± 17.4 | 24.6 ± 57.2 | 6.7 ± 3.3 | 3.8 ± 2.5 | 2.5 ± 2.2 |
| IP-10 | 601.8 ± 361.2 | 685.5 ± 652.8 | 1019.6 ± 1331.8 | 543.9 ± 83.7 | 316.6 ± 124.7 |
| MCP-1 | 261.2 ± 132.1 | 404.9 ± 369.5 | 226.4 ± 150.3 | 427.3 ± 175.7 | 491.4 ± 164.9 |
| MIP-1a | 2.4 ± 2.6 | 2.0 ± 2.4 | 2.4 ± 3.1 | 2.6 ± 1.3 | 0.0 ± 0.0 |
| MIP-1b | 23.9 ± 17.8 | 24.7 ± 12.5 | 19.1 ± 9.3 | 26.2 ± 11.4 | 20.8 ± 2.7 |
| TNFα | 10.9 ± 4.8 | 13.1 ± 6.3 | 12.4 ± 11.4 | 16.0 ± 4.8 | 12.2 ± 1.6 |
| VEGF | 36.1 ± 29.0 | 54.1 ± 76.8 | 48.3 ± 100.2 | 27.9 ± 10.8 | 16.6 ± 19.3 |
| BTLA | 1515.0 ± 1066.9 | 1772.1 ± 1416.2 | 1095.6 ± 685.2 | 1984.6 ± 1466.9 | 548.1 ± 270.4 |
| CD27 | 1685.3 ± 1462.8 | 1784.8 ± 1155.3 | 1283.8 ± 983.3 | 2387.5 ± 976.6 | 1314.5 ± 552.6 |
| CD28 | 3855.6 ± 2898.9 | 5238.6 ± 4815.2 | 9069.7 ± 20678.0 | 5640.1 ± 3279.7 | 2614.9 ± 1477.9 |
| TIM3 | 2177.8 ± 1092.2 | 3076.4 ± 1368.1 | 1908.8 ± 751.1 | 3247.4 ± 787.6 | 1793.5 ± 252.7 |
| HVEM | 1165.8 ± 454.2 | 1650.7 ± 897.7 | 1293.9 ± 1006.2 | 1485.0 ± 115.4 | 1069.0 ± 70.5 |
| CD40 | 485.0 ± 143.9 | 688.3 ± 370.5 | 588.1 ± 447.5 | 488.8 ± 116.1 | 332.3 ± 16.4 |
| GITR | 281.5 ± 310.8 | 412.7 ± 408.7 | 417.7 ± 509.5 | 346.1 ± 184.0 | 52.2 ± 60.6 |
| LAG3 | 44634.8 ± 24196.0 | 51257.0 ± 24837.0 | 55804.7 ± 23278.2 | 31734.9 ± 6971.1 | 31978.0 ± 10606.9 |
| TLR2 | 3186.3 ± 1282.4 | 3826.4 ± 2607.1 | 4951.4 ± 7797.8 | 3358.5 ± 1880.4 | 1661.1 ± 577.2 |
| GITRL | 1035.0 ± 511.2 | 1166.1 ± 696.5 | 895.9 ± 561.5 | 1166.5 ± 602.7 | 554.3 ± 155.5 |
| PD-1 | 1598.2 ± 777.8 | 1821.1 ± 1229.9 | 2978.6 ± 5506.0 | 1864.1 ± 724.7 | 587.8 ± 208.3 |
| CTLA-4 | 86.1 ± 90.2 | 103.9 ± 95.1 | 72.3 ± 65.4 | 137.6 ± 63.6 | 30.4 ± 7.0 |
| CD80 | 293.7 ± 165.5 | 350.4 ± 209.7 | 254.4 ± 131.0 | 317.5 ± 183.5 | 224.9 ± 92.5 |
| CD86 | 2003.3 ± 1220.9 | 2190.9 ± 1817.6 | 1591.0 ± 869.8 | 2219.3 ± 1300.0 | 662.8 ± 236.7 |
| PD-L1 | 319.3 ± 236.8 | 357.2 ± 234.8 | 291.2 ± 162.0 | 336.5 ± 181.3 | 124.2 ± 45.9 |
| ICOS | 4177.8 ± 2269.8 | 4482.4 ± 2686.6 | 3694.9 ± 2178.2 | 3199.1 ± 1083.1 | 1539.0 ± 585.7 |
| Arginase1 | 438.6 ± 755.0 | 443.9 ± 602.6 | 399.5 ± 637.2 | 529.1 ± 393.2 | 399.6 ± 178.4 |
| ICOSL | 3780.9 ± 3056.9 | 3792.2 ± 2377.7 | 3272.5 ± 1437.6 | 6388.8 ± 2967.1 | 5185.4 ± 1074.3 |
| CD276 | 923.8 ± 531.0 | 1150.4 ± 564.5 | 834.5 ± 426.4 | 1144.3 ± 480.4 | 1420.0 ± 448.1 |
| CD73 | 925.4 ± 970.3 | 1618.6 ± 2006.5 | 1462.5 ± 664.8 | 889.2 ± 870.7 | 550.9 ± 480.2 |
| VTN | 636.3 ± 500.6 | 804.2 ± 1344.3 | 639.3 ± 376.3 | 826.9 ± 666.2 | 934.2 ± 316.8 |
| APRIL | 375.6 ± 238.8 | 593.1 ± 687.1 | 312.0 ± 279.6 | 516.1 ± 201.7 | 307.8 ± 326.2 |
| VISTA | 45.7 ± 111.4 | 40.1 ± 52.1 | 75.6 ± 92.8 | 44.2 ± 31.4 | 34.5 ± 7.0 |
| B7-H6 | 71.9 ± 42.2 | 117.8 ± 81.2 | 94.9 ± 95.9 | 76.5 ± 62.9 | 90.0 ± 23.1 |
| Granzyme B | 8.5 ± 12.5 | 21.2 ± 78.8 | 16.3 ± 24.7 | 18.1 ± 29.3 | 15.0 ± 9.2 |
| E-cadherin | 84510.3 ± 107437.5 | 76994.1 ± 119834.5 | 66625.7 ± 49591.6 | 71242.5 ± 36336.6 | 37225.6 ± 27713.1 |
| Gal-1 | 10600.3 ± 5209.7 | 13106.9 ± 4293.6 | 9771.1 ± 2966.0 | 12342.3 ± 3163.5 | 15252.1 ± 4659.9 |
| Gal-3 | 4866.3 ± 2031.9 | 6014.1 ± 3126.8 | 5737.1 ± 3650.0 | 5652.8 ± 1760.6 | 5679.5 ± 1701.5 |
| Granulysin | 842.0 ± 445.1 | 1028.5 ± 626.0 | 653.8 ± 331.8 | 979.2 ± 492.1 | 642.4 ± 174.3 |
| IDO-1 | 161.6 ± 159.1 | 177.8 ± 188.5 | 388.2 ± 583.2 | 113.3 ± 44.5 | 65.4 ± 31.6 |
| MIC-A | 19.3 ± 13.2 | 23.5 ± 22.3 | 17.2 ± 10.9 | 28.9 ± 7.2 | 20.7 ± 5.0 |
| MIC-B | 350.3 ± 167.3 | 439.1 ± 447.6 | 375.7 ± 367.5 | 313.6 ± 237.5 | 425.6 ± 229.1 |
| BAFF | 1804.1 ± 1676.5 | 2161.8 ± 2414.3 | 916.9 ± 716.4 | 1777.5 ± 1848.3 | 703.4 ± 95.6 |
| OX-40 | 118.1 ± 64.2 | 152.7 ± 117.9 | 195.5 ± 278.7 | 204.5 ± 67.2 | 109.5 ± 36.5 |
| PVR | 64263.7 ± 42220.4 | 89382.4 ± 45399.3 | 79491.1 ± 43022.6 | 108353.6 ± 56969.0 | 74224.4 ± 38519.0 |
| Perforin | 4715.3 ± 2648.0 | 5120.1 ± 3033.6 | 4863.6 ± 2235.7 | 6052.6 ± 4226.8 | 10889.7 ± 3741.8 |
|  | 1C |  |  |  |  |
| Analyte | R | PR | SD | HPR |  |

|  |  |  |  |  |
| --- | --- | --- | --- | --- |
| **FGF-2** | 46.3 ± 37.2 | 43.6 ± 33.8 | 54.1 ± 32.0 | 34.4 ± 11.2 |
| **Eotaxin** | 107.6 ± 78.1 | 95.4 ± 58.1 | 104.1 ± 41.7 | 56.8 ± 22.8 |
| **G-CSF** | 96.4 ± 208.7 | 44.6 ± 44.9 | 30.0 ± 15.2 | 11.1 ± 9.8 |
| **GM-CSF** | 7.1 ± 8.6 | 7.5 ± 7.8 | 8.5 ± 8.9 | 8.3 ± 7.9 |
| **FKN** | **53.6 ± 26.8** | **51.2 ± 39.3** | **79.6 ± 74.2** | **45.7 ± 17.4** |
| **IFNα** | 25.6 ± 19.3 | 31.2 ± 53.4 | 22.0 ± 15.4 | 19.2 ± 3.1 |
| **IFNγ** | 7.2 ± 6.7 | 5.8 ± 3.2 | 23.9 ± 54.4 | 3.6 ± 3.4 |
| **IL-10** | 13.6 ± 27.2 | 23.9 ± 58.1 | 17.0 ± 24.3 | 10.0 ± 11.3 |
| **IL12p70** | 7.1 ± 14.3 | 4.4 ± 11.3 | 3.2 ± 1.7 | 1.6 ± 1.5 |
| **IL-15** | 0.6 ± 1.4 | 0.9 ± 1.3 | 1.2 ± 1.7 | 0.7 ± 1.2 |
| **sCD40L** | 630.8 ± 2651.4 | 406.1 ± 1089.8 | 210.3 ± 298.2 | 62.3 ± 26.5 |
| **IL-17** | 2.1 ± 2.4 | 2.4 ± 2.8 | 6.4 ± 12.5 | 0.8 ± 0.8 |
| **IL1RA** | 16.6 ± 12.3 | 43.0 ± 76.6 | 20.3 ± 17.0 | 34.7 ± 20.3 |
| **IL-1β** | 0.7 ± 1.4 | 0.9 ± 1.2 | 3.1 ± 5.8 | 0.3 ± 0.3 |
| **IL-2** | 1.4 ± 2.2 | 1.2 ± 1.7 | 0.7 ± 0.6 | 0.6 ± 0.3 |
| **IL-3** | 0.6 ± 0.7 | 0.9 ± 0.7 | 0.9 ± 0.7 | 0.5 ± 0.4 |
| **IL-5** | 0.9 ± 0.4 | 0.9 ± 1.1 | 0.5 ± 0.2 | 0.5 ± 0.1 |
| **IL-6** | 5.8 ± 10.1 | 6.4 ± 11.6 | 1.8 ± 4.0 | 1.6 ± 1.5 |
| **IL-7** | 3.2 ± 2.8 | 3.1 ± 6.5 | 1.5 ± 1.7 | 1.2 ± 1.6 |
| **IL-8** | 14.5 ± 18.5 | 28.5 ± 52.0 | 16.4 ± 14.0 | 4.8 ± 2.6 |
| **IP-10** | 951.3 ± 695.9 | 1318.2 ± 1322.4 | 1205.7 ± 1424.6 | 1295.6 ± 601.4 |
| **MCP-1** | 277.7 ± 164.6 | 444.7 ± 353.3 | 301.5 ± 301.6 | 439.8 ± 132.8 |
| **MIP-1a** | 2.6 ± 2.5 | 2.5 ± 2.6 | 4.1 ± 3.6 | 4.6 ± 5.8 |
| **MIP-1b** | 29.2 ± 24.6 | 31.5 ± 23.0 | 28.0 ± 16.3 | 34.7 ± 15.6 |
| **TNFα** | 13.6 ± 6.6 | 18.5 ± 14.2 | 19.8 ± 20.8 | 31.6 ± 21.3 |
| **VEGF** | 61.4 ± 102.9 | 52.8 ± 94.2 | 64.6 ± 121.2 | 36.4 ± 9.7 |
| **BTLA** | 1512.5 ± 1311.6 | 1887.3 ± 1406.5 | 1727.2 ± 872.0 | 1106.2 ± 444.5 |
| **CD27** | 1952.8 ± 1640.7 | 2074.5 ± 1291.8 | 1364.8 ± 1150.5 | 2680.3 ± 934.4 |
| **CD28** | 4502.8 ± 4786.3 | 5540.8 ± 5638.6 | 9454.0 ± 16205.5 | 2932.1 ± 2203.5 |
| **TIM3** | 2239.3 ± 1597.2 | 3269.4 ± 1535.6 | 2296.9 ± 886.9 | 3342.3 ± 1117.9 |
| **HVEM** | 1367.0 ± 1358.0 | 1759.0 ± 824.7 | 1473.8 ± 1387.6 | 1628.4 ± 297.2 |
| **CD40** | 516.2 ± 224.0 | 713.6 ± 287.2 | 683.3 ± 325.7 | 571.9 ± 290.2 |
| **GITR** | 335.0 ± 484.5 | 446.9 ± 435.6 | 583.6 ± 420.3 | 223.6 ± 216.9 |
| **LAG3** | 42439.5 ± 28754.3 | 53144.6 ± 26778.9 | 70465.7 ± 23905.2 | 32345.2 ± 16039.0 |
| **TLR2** | 3464.8 ± 3093.0 | 4152.5 ± 2841.1 | 5835.0 ± 6389.2 | 2468.2 ± 983.2 |
| **GITRL** | 1105.4 ± 799.2 | 1257.2 ± 790.9 | 1203.9 ± 617.3 | 799.1 ± 333.3 |
| **PD-1** | 3511.8 ± 2420.6 | 2910.8 ± 1648.4 | 5376.9 ± 6193.8 | 2382.4 ± 468.6 |
| **CTLA-4** | 99.4 ± 118.2 | 116.2 ± 108.5 | 114.1 ± 93.7 | 104.8 ± 59.8 |
| **CD80** | 330.8 ± 317.3 | 390.5 ± 282.3 | 377.7 ± 136.9 | 226.3 ± 116.8 |
| **CD86** | 1942.2 ± 1707.9 | 2269.2 ± 1461.9 | 2591.6 ± 1263.1 | 1294.2 ± 827.9 |
| **PD-L1** | 315.6 ± 319.1 | 388.0 ± 231.3 | 406.8 ± 176.5 | 216.4 ± 112.1 |
| **ICOS** | 4055.5 ± 3047.5 | 4917.4 ± 3371.1 | 5312.4 ± 2475.4 | 2615.8 ± 1157.9 |
| **Arginase1** | 666.2 ± 1457.7 | 455.1 ± 724.9 | 1359.2 ± 2865.8 | 165.6 ± 223.3 |
| **ICOSL** | 3468.4 ± 2419.7 | 3671.3 ± 2167.1 | 3369.9 ± 1241.2 | 4479.9 ± 1407.5 |
| **CD276** | 994.9 ± 571.2 | 1215.5 ± 568.8 | 886.5 ± 462.3 | 983.6 ± 542.5 |
| **CD73** | 820.7 ± 920.3 | 6576.9 ± 33384.5 | 2082.8 ± 2427.7 | 548.3 ± 775.5 |
| **VTN** | 630.4 ± 483.0 | 887.3 ± 1541.2 | 638.5 ± 396.0 | 681.2 ± 424.9 |
| **APRIL** | 623.6 ± 914.0 | 632.4 ± 616.3 | 446.5 ± 272.8 | 435.9 ± 208.6 |
| **VISTA** | 46.0 ± 81.6 | 64.8 ± 130.9 | 79.7 ± 104.0 | 43.8 ± 34.6 |
| **B7-H6** | 72.2 ± 55.4 | 128.6 ± 91.8 | 114.5 ± 125.4 | 83.0 ± 67.9 |
| **Granzyme B** | 15.4 ± 29.3 | 33.7 ± 59.6 | 21.8 ± 41.3 | 9.7 ± 10.5 |
| **E-cadherin** | 79841.8 ± 102820.9 | 73015.9 ± 89655.7 | 69720.7 ± 45131.2 | 67288.1 ± 47311.8 |
| **Gal-1** | 10904.1 ± 6870.5 | 13362.2 ± 4296.6 | 10511.1 ± 3456.1 | 11630.4 ± 3088.4 |
| **Gal-3** | 5417.5 ± 2732.0 | 6175.3 ± 2685.1 | 6828.8 ± 4745.8 | 5152.1 ± 1429.9 |
| **Granulysin** | 1055.6 ± 983.2 | 1350.7 ± 904.6 | 908.7 ± 584.5 | 944.6 ± 324.4 |
| **IDO-1** | 322.8 ± 584.5 | 214.2 ± 243.7 | 494.7 ± 772.7 | 142.2 ± 65.6 |
| **MIC-A** | 20.4 ± 14.3 | 29.0 ± 38.7 | 18.7 ± 11.7 | 31.9 ± 15.9 |
| **MIC-B** | 330.5 ± 123.6 | 544.3 ± 825.6 | 379.2 ± 323.4 | 346.8 ± 294.0 |
| **BAFF** | 2319.9 ± 3341.6 | 3046.8 ± 2749.2 | 2074.1 ± 2398.8 | 2157.1 ± 1061.0 |
| **OX-40** | 180.1 ± 151.4 | 186.5 ± 83.0 | 272.9 ± 432.5 | 313.5 ± 159.1 |
| **PVR** | 66329.9 ± 39760.1 | 93820.7 ± 54476.9 | 81003.1 ± 47797.7 | 95843.9 ± 37313.2 |
| **Perforin** | 5521.4 ± 3508.9 | 6926.9 ± 4620.1 | 5881.6 ± 4255.8 | 6732.8 ± 5220.3 |

## Data Availability

All data produced in the present study are available upon reasonable request to the authors.

## Acknowledgments

We sincerely acknowledge all the patients and families that generously agreed to participate in this study. We are also thankful to the nursing staff of the Medical Oncology Day Care at University Hospital of Navarre who kindly provided the clinical samples.

## Funding

This research was supported by:

The Spanish Association against Cancer (AECC), PROYE16001ESCO, IDEAS211016AJON to (DE, GK, RV)

Instituto de Salud Carlos III (ISCIII)-FEDER Project grants FIS PI17/02119, FIS PI20/00010, FIS PI20/00419; COV20/00000, and TRANSPOCART ICI19/00069 granted to (DE, GK).

Biomedicine Project Grant from the Department of Health of the Government of Navarre- FEDER funds (BMED 050-2019, 51-2021) granted to (DE).

Strategic projects from the Department of Industry, Government of Navarre (AGATA, Ref. 0011-1411-2020-000013; LINTERNA, Ref. 0011-1411-2020-000033; DESCARTHES, 0011-1411-2019-000058) granted to (DE, GK, RV).

European Project Horizon 2020 Improved Vaccination for Older Adults (ISOLDA) under the grant agreement ID: 848166) granted to (DE, GK).

## Author contributions

A.B., D.A., G.K., D.E., R.P., R.V., L.M. and J.J.L. were responsible for the study conception and design. G.F., H.A., M.M., I.M. and R.V. were responsible for patient recruitment and clinical samples supply. A.B., E.B., D.A. and A.R. carried out the *in vivo* experiments. A.B., P.M., P.R., E.B., L.F. and G.K. performed cell lines engineering. A.B., G.F., D.A. and B.T. carried out luminex assays. A.B. was responsible for flow cytometric data acquisition and analysis. G.F. and H.A. compiled and analyzed patient medical histories. L.C., S.P. and G.F. were responsible for bioinformatics analysis. A.B., M.Z., M.G., H.A., E.B., L.C., M.E. and G.K. were responsible for clinical samples processing. A.B., G.F., D.A., H.A., M.Z., L.C., S.P., D.E., R.P. and G.K. performed the analysis and interpretation of the data. G.F., S.P. and D.E. performed the statistical analysis. A.B., D.A., R.P., D.E. and G.K. wrote the manuscript. All authors critically reviewed the manuscript and approved its final version.

## Data and materials availability

All data are available in the main text or the supplementary materials.

